# Replacing animal-sourced with plant-sourced proteins alters gut microbiota and biomarkers associated with colorectal cancer risk in healthy adults: A randomized feeding study

**DOI:** 10.64898/2026.10.13.26365402

**Authors:** Ching Jian, Tiina Pellinen, Antti Karkman, Mikko Neuvonen, Mikko Niemi, Anne Salonen, Anne-Maria Pajari

**Author notes:** Corresponding authors: Ching Jian & Anne-Maria Pajari.

## Abstract

Epidemiological studies associate replacing animal-sourced with plant-sourced protein with lower colorectal cancer (CRC) risk, but controlled feeding studies have rarely tested how this substitution alters the gut microbiota and CRC-related biomarkers. In a 12-week randomized controlled feeding study, 136 healthy adults consumed diets providing 30 energy percent (E%), 50 E%, or 70 E% of protein from plant sources. Change in gut microbiota composition was the primary outcome; stool characteristics, fecal *N*-nitroso compounds (NOCs), and fecal and plasma bile acids (BAs) were secondary outcomes. The 70% plant-protein diet significantly restructured the gut microbiota, increasing fiber-associated *Lachnospiraceae* and *Roseburia* while reducing *Methanobrevibacter* and several asaccharolytic taxa. Microbial functional potential shifted toward plant-carbohydrate degradation, with lower amino acid degradation and methanogenesis. Fecal pH decreased, stool characteristics suggested faster colonic transit, and potentially carcinogenic total and haem-derived NOCs declined progressively with plant-protein replacement. The 70% plant-protein diet also reduced fecal secondary and taurine-conjugated BAs, including the CRC-associated BAs DCA and LCA. Plasma glucuronidated BAs increased, expanding the circulating pool of readily excreted BA conjugates. Higher insoluble fiber intake was associated with greater microbiota restructuring and lower fecal NOCs. Methanogen carriers had higher baseline fecal NOCs and features of slower colonic transit, and showed weaker microbiota and less favorable BA responses to the 70% plant-protein diet. Baseline methanogen carriage thus could be exploited for participant stratification in future dietary interventions. ClinicalTrials.gov: NCT03206827.

## Introduction

Dietary protein intake in many Western countries remains dominated by animal sources, which account for approximately 60-70% of total protein intake (Halkjaer et al. 2009). Environmental concerns and the burden of chronic diseases have increased interest in replacing part of animal protein with plant-sourced protein (Davis et al. 2026). Colorectal cancer (CRC) is the third most commonly diagnosed cancer and the second leading cause of cancer death worldwide (Siegel et al. 2023). Epidemiological studies suggest that replacing red and processed meat, or animal protein more broadly, with plant-sourced protein is associated with lower CRC risk (Budhathoki et al. 2019; Sieri et al. 2022; Tammi et al. 2024). In a two-week dietary exchange between African American and rural South African adults, switching between a Western-style, animal-protein-dominant diet and an African-style, plant-protein-dominant diet produced reciprocal changes in colonic mucosal proliferation, inflammatory markers, and gut microbial metabolism associated with CRC risk (O’Keefe et al. 2015). However, this intervention was designed to compare contrasting dietary patterns, not a systematic replacement of animal-sourced with plant-sourced protein. Protein intake also differed substantially between the intervention diets (27 vs. 15 E%), making it difficult to attribute the observed effects to protein source.

The gut microbiota is one potential link between diet and colorectal carcinogenesis. Metagenomic analyses across multiple cohorts have identified increased microbial capacity for protein and mucin degradation and reduced carbohydrate degradation in CRC-associated microbiomes (Wirbel et al. 2019). Elevated proteolytic fermentation in the colon can increase the production of potentially harmful microbial metabolites such as *p*-cresol and hydrogen sulfide (H S), while enhanced mucin degradation may erode the colonic mucus barrier (Procházková et al. 2023). Diet-derived substrate availability has an impact on these microbial metabolic processes. Plant proteins generally have lower ileal digestibility than animal proteins, which could increase the amount of protein reaching the colon (Bartlett and Kleiner 2022; Davis et al. 2026). However, plant-protein whole foods also supply dietary fiber and other fermentable carbohydrates that may alter microbial substrate use and reduce reliance on proteolytic fermentation (Davis et al. 2026). The net effect of replacing animal-sourced with plant-sourced protein on colonic microbial metabolism is therefore difficult to infer from protein digestibility alone, and controlled human studies are lacking (Davis et al. 2026).

Diet can modify the physicochemical and chemical environment of the colon in ways relevant to colorectal carcinogenesis, as reflected in stool characteristics such as fecal weight, pH, and consistency. Higher fecal weight and lower fecal pH have been associated with lower CRC risk, whereas harder stools reflect slower colonic transit and prolonged contact between the colonic mucosa and fecal contents (Davies et al. 1986; Cummings et al. 1992; Vandeputte et al. 2016; Yamamura et al. 2023; Procházková et al. 2024). These contents include potentially carcinogenic compounds such as *N*-nitroso compounds (NOCs), some of which can form DNA-alkylating intermediates and have been proposed as one mechanism linking red and processed meat intake with CRC (Bingham et al. 2002; Seiwert et al. 2021; Ruiz-Saavedra et al. 2024). Also, bile acids (BAs) are extensively transformed by gut microbes in the colon, generating secondary BAs that at elevated concentrations can disrupt epithelial cell membranes and induce genotoxic stress (Jia et al. 2018).

Whole-food feeding studies that replace animal-sourced with plant-sourced protein remain very rare (Courtney et al. 2026), since providing and controlling most of participants’ diets is logistically demanding. Here, we conducted a 12-week randomized controlled isocaloric feeding study with 136 healthy working-age Finnish adults assigned to diets providing 30 E%, 50 E%, or 70 E% of total protein from plant sources. The primary outcome was change in gut microbiota composition; secondary outcomes included fecal and plasma CRC risk-related biomarkers (stool characteristics, fecal NOCs, fecal and plasma BAs) measured at baseline and week 12. Fecal microbiota was profiled by 16S rRNA gene sequencing in the full cohort and by shotgun metagenomic sequencing in the two most contrasting diet groups. Fecal NOCs were quantified by selective denitrosation, and fecal and plasma BAs by targeted metabolomics. Together with detailed dietary records and measurements of stool characteristics, these analyses provided an integrated assessment of the colonic adaptation to graded replacement of animal-sourced protein with plant-sourced protein within a realistic Western dietary setting.

## Methods

### Study design and participants

The ScenoProt intervention study was a single-site, three-arm, parallel-group randomized controlled feeding study conducted at the Department of Food and Nutrition, University of Helsinki, Finland, to investigate the effects of varying proportions of animal- and plant-based protein on nutritional intake, metabolic, and gut health. The primary outcome as listed on ClinicalTrials.gov was the change in gut microbiota composition within each arm from baseline (week 0) to end of intervention (week 12). Changes in stool characteristics, fecal NOCs, fecal and plasma BAs measured at baseline and at week 12 were secondary outcomes. Screening was conducted from December 2016 to March 2017, and follow-up ended in June 2017. The study protocol and findings from the nutritional analyses have been published (Päivärinta et al. 2020; Pellinen et al. 2022).

Community volunteers were recruited through newspaper advertisements, social media, and university mailing lists. Eligible participants were healthy omnivorous adults aged 20-69 years with a BMI of 18.5-35.0 kg/m². Exclusion criteria included fasting plasma glucose >6.9 mmol/L, total cholesterol >6.5 mmol/L, medication for diabetes or hypercholesterolemia, gastrointestinal, endocrine, renal, or liver diseases, eating disorders, recent antibiotic use (past three months), food allergies, smoking, pregnancy, lactation, or participation in extreme sports. Participants stopped dietary supplements, herbal remedies, and natural products at least two weeks before the intervention and were asked to maintain their usual physical activity and medical care.

The study was approved by the Coordinating Ethics Committee of the Hospital District of Helsinki and Uusimaa (1651/2016), and all participants provided written informed consent. The study was conducted in accordance with the Declaration of Helsinki. This clinical study was registered at ClinicalTrials.gov (NCT03206827). There was no formal patient or public involvement in the design, conduct, analysis, or reporting of the study.

### Sample size estimation and randomization

The sample size was calculated to detect differences in haem-derived *N*-nitroso compound concentrations in feces, as this was expected to be the most variable outcome. Power calculations were based on data from a previous 4-week intervention, where haem-derived *N*-nitroso compounds measured 2.875 ± 2.800 pmol/mg feces before the intervention and 2.245 ± 1.125 pmol/mg feces post-intervention. With a two-sided α of 0.05 and 80% power, the target sample size was 50 participants per group. Recruitment ceased when the scheduled intervention cohorts had been filled, and the trial was not stopped early.

The intervention was staggered over 11 weeks, ensuring equal distribution of participants across diet groups. Within age and sex strata, participants were assigned with equal probability to the Animal, 50/50, or Plant group, targeting similar age and sex distributions and a 1:1:1 allocation ratio. The principal investigator generated the allocation sequence and randomly assigned the participants into intervention groups within a similar deviation of age and gender. Blinding was not possible due to the nature of the intervention However, participants were unaware of the exact animal-to-plant protein ratio in their assigned diet, as color-coded meal plans were used instead of explicit diet labels (Päivärinta et al. 2020).

### Intervention diets

Eligible participants were randomly assigned in a 1:1:1 ratio to one of three diet groups: (1) Animal: A diet representing an average Finnish diet, with 70% of protein from animal sources (red and white meat, fish, eggs, dairy) and 30% from plant sources. (2) 50/50: A mixed-protein diet with equal proportions (50%) of animal and plant proteins, limiting red and processed meat intake to ≤500 g per week. (3) Plant: A predominantly plant-based diet with 70% of protein from plant sources and 30% from animal sources. All diets were designed to provide 17% of total energy intake (E%) from protein while differing in protein sources. In the Animal group, protein intake came primarily from red meat, poultry, dairy, and fish, while in the 50/50 and Plant groups, animal proteins were progressively replaced with plant-based sources, including legumes (peas, lentils, faba beans, chickpeas), whole grains, nuts, seeds, and tofu. Across all groups, fish intake (250 g/week) and egg consumption (four eggs per week) were standardized. To ensure compliance, 80% of daily energy intake was provided by the study in the form of protein-rich foods (e.g., meat, poultry, plant-based products, nuts, and pulses). Dairy and eggs were self-selected according to dietary instructions. Participants were allowed to consume habitual amounts of low-protein foods, including fruits, vegetables, confectionery, and beverages. Energy contents of the intervention diets were based on average energy consumption of 8400 kJ (2000 kcal) but the participants were advised to maintain a stable weight and to eat according to their appetite.

To facilitate dietary adaptation and reduce gastrointestinal discomfort, Plant-group participants followed the 50/50 diet during the first week before moving to the Plant diet. Participants visited the research centre weekly to receive study foods, be weighed by a nutritionist, and receive individualized dietary guidance. Participants were asked to avoid nutritional supplements and natural products and to maintain their usual physical activity. Further details about intervention diets are reported elsewhere (Päivärinta et al. 2020).

### Assessment of dietary intake

Dietary intake was assessed using 4-day food records, completed at baseline and during the final week of the intervention. Additionally, participants submitted one-day food records every third week to monitor adherence. Participants received detailed written and verbal instructions on accurately recording food intake, including estimating portion sizes using kitchen scales, measuring tools, and food packaging labels. The 4-day records included three weekdays and one weekend day to account for variations in dietary habits.

At baseline, 118 participants submitted complete 4-day food records, while 11 recorded three days, and one recorded two days. At the end of the intervention, 117 participants provided 4-day records, while 16 recorded three days, one recorded two days, and one recorded one day. Missing records (N = 4) were replaced with additional food records collected during the intervention. Incomplete or inconsistent records at baseline and endpoint were excluded from analysis. Some participants (N = 4 at baseline; N = 1 at endpoint) recorded more than four days, but only the first four were analyzed.

Food records were processed using AivoDiet software (version 2.2.0.1, Aivo Oy, Turku, Finland), linked to the Fineli^®^ Food Composition Database (Release 16, Finnish Institute for Health and Welfare, 2013). If necessary, new food items were added to the database. Recorded meals were matched with existing database recipes, or new recipes were created when needed. Nutrient intakes were calculated as daily means or medians over the recorded period.

To classify dietary intake, all food items were categorized into 14 food groups following a modified version of the Fineli^®^ classification system. An additional category for plant-based dairy alternatives was introduced. Plant-protein-based dishes were grouped under vegetable or plant-based meal categories. The contribution of different food categories to total energy and nutrient intake was analyzed, distinguishing between animal- and plant-based sources where applicable. More details regarding dietary assessments and findings are available in our previous publications (Päivärinta et al. 2020; Pellinen et al. 2022). To capture changes in overall dietary composition during the intervention, Bray–Curtis dissimilarity between baseline and endpoint was calculated from intake of the 14 food groups (i.e., food distance) for each participant (Troci et al. 2023; Yamamoto-Wada et al. 2025).

### Blood and fecal sample collection and processing

At the beginning and end of the 12-week intervention, venous blood samples were collected after 10-12 hours of fasting, with 5 mL drawn into Aprotinin K3EDTA tubes. Samples were immediately centrifuged at 3000 rpm and 4°C for 15 minutes to separate plasma, then aliquoted into cryotubes and stored at -70°C until analysis. Fecal samples were collected as a complete bowel movement, immediately frozen at -20°C at the participant’s home, and transported to the university laboratory, where they were stored at −70°C within 24 hours of defecation. During processing, samples were thawed at 4°C, and wet weight was recorded to the nearest 0.01 g after subtracting collection material weight. Fecal pH was measured using a Symphony SB70P pH meter (VWR International, LLC., Radnor, PA, USA). Stool consistency was assessed using the Bristol Stool Scale (BSS), and aliquots were taken from the middle portion for microbiota and metabolome profiling and *N*-nitroso compound quantification.

### Quantification of fecal total NOCs and haem-derived NOCs

Fecal homogenates were prepared (1:5 dilution in Milli-Q water) and homogenized with an Ultra Turrax. Total and haem-derived NOCs were analyzed using selective denitrosation and chemiluminescence-based detection with an Ecomedics CLD 88 Exhalyzer, as previously described in detail (Ruiz-Saavedra et al. 2024). A preservation solution prevented artificial nitrosation, and acid sulphanilamide (SA) solution removed nitrite. Haem-derived NOCs were determined indirectly using HgCl and K Fe(CN). Samples were incubated with reagents before injection into a triiodine mixture at 60°C, releasing NO for detection. Each sample was injected twice with a 5-10 min interval. Signal detection used a PowerChrom 280 system, with quantification based on sodium nitrite calibration curves (5-950 pmol, *R*² > 0.96). The quantification limit was 2.5 pmol, and haem-derived NOCs were calculated by subtracting nonheme values from total NOCs.

### Bile acid profiling by UHPLC-MS/MS

Cholic acid (CA), chenodeoxycholic acid (CDCA), deoxycholic acid (DCA), glycholic acid (GCA), glycodeoxycholic acid (GDCA), glycochenodeoxycholic acid (GCDCA), glycolithocholic acid (GLCA), glycoursodeoxycholic acid (GUDCA), hyodeoxycholic acid (HDCA), lithocholic acid (LCA), taurocholic acid (TCA), taurochenodeoxycholic acid (TCDCA), taurodeoxycholic acid (TDCA), taurolithocholic acid (TLCA), tauroursodeoxycholic acid (TUDCA) and ursodeoxycholic acid (UDCA) were purchased from Cayman Chemical (MI, USA). Glycochenodeoxy cholic acid 3-O-glucuronide (GCDCA-3G), glycodeoxycholic acid 3-O-glucuronide (GDCA-3G), glycochenodeoxy 3-sulfate (GCDCA-3S) and glycodeoxycholic acid 3-sulfate (GDCA-3S) were from Toronto Research Chemicals (Toronto, Canada). The bile acid (BA) concentrations were assayed from plasma and fecal extracts at the Department of Clinical Pharmacology, University of Helsinki, Helsinki, Finland.

Plasma samples along with the reference calibration standards (0.001-5 µM) and quality controls were thawed in room temperature and prepared by protein precipitation. A 100 µL aliquot of plasma was combined with 400 µL of acetonitrile containing stable isotope-labeled internal standards. This mixture was processed on a 96-well Impact filter plate (Phenomenex, Torrance, CA) utilizing a Freedom EVO automated liquid handling system (Tecan Group, Männedorf, Switzerland). Following mixing, the sample mixture was incubated for 15 minutes at 4°C, after which the supernatant was separated. The extracted sample was then dried using a GeneVac centrifugal evaporator (Thermo Fisher Scientific, Waltham, MA) and subsequently reconstituted in 50 µL of 50% methanol. The fecal BAs were extracted using alkaline hydrolysis and mechanical lysis of lyophilized fecal samples, followed by cartridge-based solid phase extraction (modified from (Humbert et al. 2012)).

The measurement of individual BAs was performed on HSS T3 analytical column (2.1x100 mm I.D.; 1.8 µm particle size) (Waters, Milford, MA, USA) using a SCIEX 6500QTrap UHPLC-MS/MS system (ABSciex, Toronto, ON, Canada) as previously described with some modifications (Xiang et al. 2010). The mobile phase was a mixture of 0.1% formic acid (solvent A) and 0.1 % formic acid in methanol (solvent B). The mobile phase gradient was structured as follows: an isocratic hold of 45% B for 1.5 minutes, followed by a linear gradient increase from 45% B to 95% B over 8.5 minutes, then a 2-minute hold at 95% B before returning to the initial condition of 45% B. The column was maintained at a temperature of 40°C, with a flow rate of 0.5 mL/min. The mass spectrometer operated in negative ion mode using multiple reaction monitoring (MRM), with an electrospray ion source temperature set at 450°C. Peak integration was performed with Analyst 1.7 software using the corresponding isotope labeled internal standards for each analyte except for GLCA, for which CDCA-D4 served as the internal standard. The plasma CA, TCA, TCDCA, TDCA, GLCA, TLCA, GUDCA, TUDCA, GCDCA-3G, GDCA-3G, GCDCA-3S, GDCA-3S had a lower limit of quantification (LLOQ) of 0.001 µM. Meanwhile, GCA, UDCA, CDCA, DCA and GDCA had a LLOQ of 0.005 µM, GCDCA 0.025 µM and HDCA 0.05 µM. For the fecal BAs, expressed as arbitrary units relative to their corresponding internal standards, a signal-to-noise (S/N) ratio of 7 was used as the lowest measurable level. The between-day precisions for all BA quality controls (0.02 µM and 2.0 µM) were below 12% and accuracies within 13%.

Individual BAs were further summed into non-mutually exclusive groups according to metabolic origin and chemical modification. Primary BAs comprised CA, CDCA, GCA, GCDCA, TCA, and TCDCA, whereas secondary BAs comprised DCA, LCA, UDCA, HDCA, GDCA, GLCA, GUDCA, TDCA, TLCA, and TUDCA, distinguishing BAs synthesized in the liver from those formed through intestinal microbial transformation (Wahlström et al. 2016). Conjugation status was assessed separately as unconjugated BAs (CA, CDCA, DCA, LCA, UDCA, and HDCA), glycine-conjugated BAs (GCA, GCDCA, GDCA, GLCA, and GUDCA), and taurine-conjugated BAs (TCA, TCDCA, TDCA, TLCA, and TUDCA). The 12α-hydroxylated BA pool comprised CA, GCA, TCA, DCA, GDCA, and TDCA, representing the CA/DCA lineage of BA metabolism (Haeusler et al. 2013). Sulfated BAs (GCDCA-3S and GDCA-3S) and glucuronidated BAs (GCDCA-3G and GDCA-3G) were analyzed separately as phase II conjugates involved in BA detoxification and elimination (Alnouti 2009; Perreault et al. 2013).

### DNA extraction and sequencing for gut microbiota profiling

Microbial DNA was extracted from approximately 250 mg of fecal samples using a modified repeated bead beating method, optimized for efficient DNA extraction from both Gram-positive and Gram-negative bacteria (Salonen et al. 2010). Briefly, fecal material was suspended in 0.5 ml of sterile ice-cold PBS, and 250 μl of fecal suspension was combined with 340 μl of RBB lysis buffer (500 mM NaCl, 50 mM Tris-HCl (pH 8.0), 50 mM EDTA, 4% SDS) in a bead-beating tube from the Ambion MagMAX™ Total Nucleic Acid Isolation Kit (Life Technologies). Following bead beating, 200 μl of the supernatant was used for DNA extraction with the KingFisher™ Flex automated purification system (Thermo Fisher Scientific) using the MagMAX™ Pathogen High Volume protocol. DNA concentration was determined using the Quanti-iT™ PicoGreen dsDNA Assay (Invitrogen).

For 16S rRNA gene amplicon sequencing, the hypervariable V3-V4 regions of the 16S rRNA gene were amplified using the 341F/785R primer pair and sequenced on the Illumina MiSeq platform following the manufacturer’s protocol, with a slight modification where TruSeq-tailed one-step amplification (Raju et al. 2018) was employed for library preparation as detailed previously (Luukkonen et al. 2018).

Shotgun metagenomic sequencing was performed on the same DNA extracts for samples in the Animal and Plant groups. Library preparation was carried out using the NEBNext Ultra II DNA Library Prep Kit for Illumina (Illumina, USA). The libraries were pooled and sequenced on the NovaSeq 6000 S4 PE150 platform (2 × 150 bp paired-end sequencing).

### Bioinformatics analysis

16S rRNA gene amplicon sequencing generated an average of 58,300 ± 22,100 sequences per sample. Demultiplexed reads were processed using QIIME2, with residual primers and adapters removed using the *cutadapt trim-paired* plugin. The *dada2 denoise-paired* pipeline was applied to truncate low-quality bases, join paired-end reads, remove chimeras, and generate amplicon sequence variants (ASVs). Taxonomic classification was performed using the SILVA database (v132), where the hypervariable V3-V4 region was extracted with the *feature-classifier extract-reads* function and used to train the Naïve Bayes classifier via *feature-classifier fit-classifier-naive-bayes*. Microbiota α-diversity (observed richness, Shannon index, and inverse Simpson’s index) and Bray-Curtis dissimilarity were calculated after rarefying bacterial sequences to 7,200 per sample, ensuring all samples were retained for further analyses. To minimize spurious taxa, microbial taxa with a relative abundance < 0.25% in < 10% of samples were excluded from downstream statistical analysis (Reitmeier et al. 2021).

Shotgun metagenomic sequencing generated an average of 54 ± 16.8 million sequences per sample. Raw reads underwent quality control using FastQC, followed by adapter removal with cutadapt v.3.5 (Martin 2011). Host sequences were removed by mapping the reads against the human genome (GRCh38) with Bowtie2 v.2.4.4 (Langmead and Salzberg 2012), microbial taxonomic profiles were generated using MetaPhlAn 4.0 (Blanco-Míguez et al. 2023) with default parameters and using the database mpa_vJan21_CHOCOPhlAnSGB_202103. For functional profiling, high-quality paired-end reads were co-assembled per subject (baseline and end-of-intervention samples) with spades v. 3.15.5 (Prjibelski et al. 2020). Genes were predicted using prodigal v.2.6.3 (Hyatt et al. 2010), and a non-redundant gene catalog of 5 million microbial genes was constructed using Cd-hit v4.8.1 with a sequence identity threshold of 95%. Gene abundance was estimated by mappping the reads against the gene catalogue using Bowtie2. Non-redundant gene sets were mapped to the EggNOG v5.0 orthologous groups database using EggNOG-mapper v2.0.1 in Diamond mode to obtain Kyoto Encyclopedia of Genes and Genomes (KEGG) orthology (KO) and carbohydrate-active enzyme (CAZyme) annotations. Metagenomic functional profiles were determined using Gut Metabolic Modules (GMMs), a set of 103 conserved metabolic pathways defined by KO identifiers. Additional targeted functional modules (e.g., flagellar assembly) were manually curated based on KO annotations (Kanehisa et al. 2016). CRC-relevant virulence factors (*bai*, *pks*, *fadA*, and *bft*) were identified using their pre-constructed Hidden Markov Models (HMMs; available in (Wirbel et al. 2019)) via HMMSearch v3.3.0. CAZymes involved in carbohydrate metabolism were grouped according to the classification provided by Li et al. (Li F et al. 2025). For downstream analysis, microbial species were retained if they had a minimum relative abundance of 0.01% in at least 10% of samples, while gene content was included if its relative abundance exceeded 0.001% in at least 10% of samples.

To characterize the resistomes, the reads generated by shotgun metagenomic sequencing were mapped against the ResFinder4 database using Bowtie2 with default settings. ResFinder4 includes clinically relevant and taxa-nonspecific antimicrobial resistance genes (ARGs) that can be vertically and horizontally transferred (acquired resistance) (Bortolaia et al. 2020). The α-diversity indices and Bray-Curtis dissimilarity of ARGs were computed similarly to ASV-based diversity indices.

Species replication rates were predicted from metagenome-assembled genomes constructed with anvi’o (Eren et al. 2021) and Semibin2 (Pan et al. 2022) using the growth rate index (GRiD) algorithm (v1.3) with default settings, which leverages coverage differences over a microbe’s genome to infer microbial growth rate as a proxy for metabolic activity in a community (Emiola and Oh 2018). Genomes with coverage < 1× and species heterogeneity > 0.3 were discarded to remove spurious growth rate estimates (Emiola and Oh 2018).

### Statistical analysis

All statistical analyses were performed in R (v4.4.1) unless otherwise stated. Analyses included the 136 participants who completed the 12-week intervention, with available data used for each measurement and no imputation of missing values. Data distributions were examined using histograms, boxplots, and quantile–quantile plots. Figures were generated using*ggplot2* (v3.5.1), *ggpubr* (v0.6.0), *cowplot* (v1.1.3), *microViz* (v0.12.4), and *pheatmap* (v1.0.12), alongside other R packages specified below.

To remain consistent with previous reports from the ScenoProt intervention (Pajari et al. 2019; Päivärinta et al. 2020; Pellinen et al. 2022), between-group differences in stool characteristics, fecal total and haem-derived NOCs, and microbiota α-diversity at the end of the intervention were analyzed by analysis of covariance (ANCOVA), adjusting for the corresponding baseline value, followed by Bonferroni-adjusted pairwise comparisons. Within-individual changes in food-group intake and microbiota composition were calculated as Bray-Curtis dissimilarities between baseline and post-intervention profiles, and differences in these distances between diet groups were tested using Wilcoxon rank-sum tests. Changes in fiber intake were assessed within groups using paired tests and between groups using linear mixed-effects models (LMEs). For analyses not subjected to multiple-testing correction, *p* < 0.05 was considered statistically significant and *p* < 0.10 was reported as a trend where indicated.

Differences in overall microbiota composition were assessed using PERMANOVA based on Bray-Curtis dissimilarity (*adonis*2, *vegan* v2.6.10), with 999 permutations (Oksanen et al. 2013). Pre-post changes were tested separately within each diet group. PERMANOVA was also used to compare baseline microbiota composition between methanogen carriers and non-carriers and to assess pre-post microbiota changes after the Plant diet separately in carriers and non-carriers.

Individual BA measurements and microbial taxonomic and functional features were analyzed separately for between-group and within-group changes. Between-group differences in pre-post change were tested using LMEs implemented in *MaAsLin2* (v1.18.0) (Mallick et al. 2021). Participant ID was included as a random effect, and intervention group, time, and the group × time interaction were included as fixed effects. The interaction term tested whether the change from baseline differed between diet groups. Within-group pre-post changes were assessed using Wilcoxon signed-rank tests. BA concentrations were rank-based inverse-normal transformed before analysis, whereas microbial relative abundances were log-transformed to improve normality and stabilize variance (Chen et al. 2020; Gao et al. 2024; Li F et al. 2025).

Baseline differences between methanogen carriers and non-carriers were assessed using two-sample *t*-tests or Wilcoxon rank-sum tests according to data distribution. BA responses to the Plant diet were compared between carriers and non-carriers using LMEs with participant ID as a random effect and carriage status, time, and their interaction as fixed effects. The carriage status × time interaction tested whether the pre-post change differed between carriers and non-carriers.

Mantel tests were used to examine associations among dietary, microbiota, metagenomic, and BA datasets (*vegan* v2.6.10) (Oksanen et al. 2013). Bray-Curtis dissimilarity was used for dietary and microbiota data and Euclidean distance for BA data, as described previously (Jacobs et al. 2023). Spearman correlations were used to examine associations among changes in bacterial genera, dietary intake, fecal NOCs, stool characteristics, and selected microbial functions over the intervention. Chord diagrams were generated using *circlize* (v0.4.16) (Gu et al. 2014).

For analyses involving multiple BAs, bacterial taxa, or microbial functions, *p* values were adjusted using the Benjamini-Hochberg method. Adjusted *p* values are reported as *q* values, with *q* < 0.2 considered significant for these analyses, in line with previous microbiome studies of similar sample size (Jeyaram et al. 2025; Wu et al. 2025).

## Results

### Graded replacement of animal-sourced with plant-sourced protein increases total and insoluble fiber intake

ScenoProt was a 12-week randomized whole-food feeding study in which animal-sourced protein was progressively and in isoenergetic means replaced with plant-sourced protein (Fig. 1A). The study protocol and nutritional outcomes have been described previously; no serious intervention-related adverse events occurred (Päivärinta et al. 2020; Pellinen et al. 2022). Of 543 people who responded to recruitment advertisements, 179 completed eligibility screening. Nine did not meet the eligibility criteria and 24 declined participation. Of the remaining 146, one withdrew for personal reasons before randomization and 145 began the intervention (113 women, 32 men). Participants consumed an Animal diet (70 E% animal, 30 E% plant protein), a 50/50 diet (50 E% animal, 50 E% plant protein; red and processed meat ≤500 g/week), or a Plant diet (30 E% animal, 70 E% plant protein). Fish and egg intake was similar across groups. Study foods given to participants provided approximately 80% of daily energy intake and covered most of their protein intake, while participants obtained fats and low-protein foods themselves. Nine participants (6.2%) discontinued the intervention: two in the Animal group, two in the 50/50 group, and five in the Plant group. Reasons were inability to follow the diet (N = 5), an unmet inclusion criterion identified after enrollment (N = 1), unrelated illness (N = 2), and medication use (N = 1). The final analyses included 136 participants (107 women, 29 men; mean age 48 years; mean BMI 24.7 kg/m²). Baseline dietary intake and participant characteristics were similar across groups (Table 1). Before the intervention, 62.4% of protein intake came from animal sources and mean fiber intake was 28.7 g/day (Päivärinta et al. 2020). Dietary compliance, assessed from food records and 24-h urinary nitrogen excretion, was high (Päivärinta et al. 2020). Energy, fat, and carbohydrate intake remained similar among groups during the intervention, although protein intake was slightly higher in the Animal (∼18 E%) and 50/50 (∼17 E%) groups than in the Plant group (∼15 E%) (Päivärinta et al. 2020).

**Figure 1.**
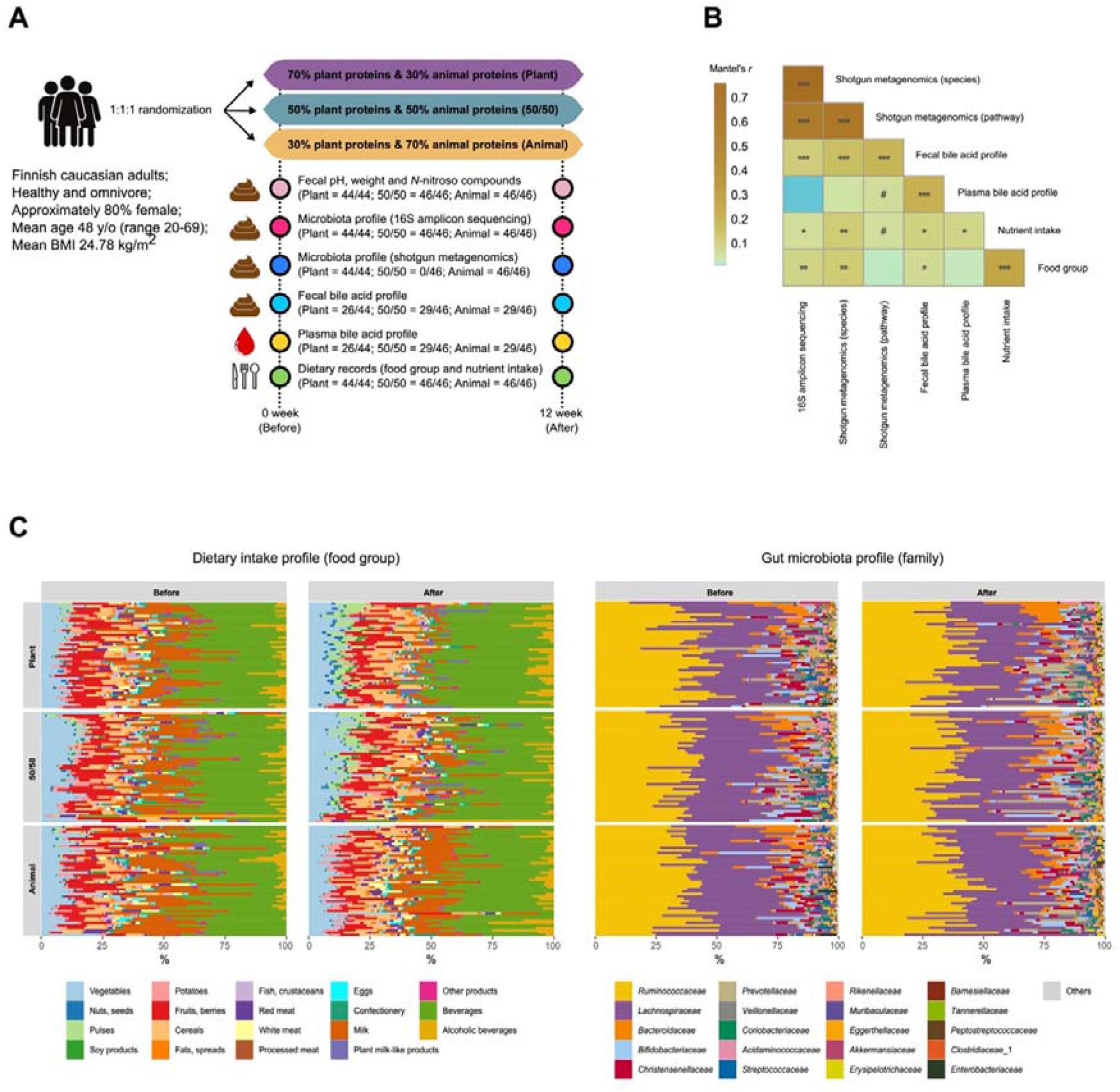
Study design, cross-dataset associations, and dietary and gut microbiota profiles during the ScenoProt intervention. **(A)** Schematic of the 12-week randomized intervention, showing the three diets and the number of participants with paired baseline and post-intervention data available for each analysis. **(B)** Pairwise Mantel correlations among dietary, microbiota, metagenomic, and bile acid datasets. Colors indicate Mantel’s *r*; statistical significance is denoted by #*p* < 0.1, \**p* < 0.05, \*\**p* < 0.01, \*\*\**p* < 0.001. **(C)** Food-group intake profiles and family-level gut microbiota composition at baseline and at the end of the intervention in the Animal, 50/50, and Plant groups. Each horizontal bar represents one participant. Baseline and end-of-intervention measurements are denoted as “Before” and “After,” respectively.

**Table 1.** Baseline characteristics of healthy adult participants assigned to the 12-week intervention diets differing in the proportions of animal- and plant-sourced protein.

|  | **Animal (N = 46)** | **50/50 (N = 46)** | **Plant (N = 44)** |
| --- | --- | --- | --- |
| Age (years) | 47.57 ± 14.50 | 47.20 ± 14.74 | 48.70 ± 14.01 |
| Sex (%female) | 80 % | 78 % | 77 % |
| BMI (kg/m<sup>2</sup>) | 24.71 ± 4.13 | 24.42 ± 3.92 | 25.23 ± 3.72 |
| Physical activity (times per week) | 3.23 ± 1.64 | 3.28 ± 1.70 | 3.39 ± 1.71 |
| Fecal pH | 6.93 ± 0.36 | 6.82 ± 0.37 | 6.91 ± 0.33 |
| Fecal wet weight (g) | 396.61 ± 184.42 | 389.13 ± 194.04 | 400.60 ± 204.30 |
| Stool consistency (BSS) | 3.85 ± 0.67 | 3.61 ± 0.58 | 3.61 ± 0.72 |
| Total fecal NOCs (pmol/mg) | 4.29 ± 3.90 | 4.24 ± 3.30 | 4.53 ± 3.04 |
| Haem-derived fecal NOCs (pmol/mg) | 2.85 ± 3.28 | 2.59 ± 2.51 | 2.88 ± 2.41 |

The change in protein source resulted in marked changes in the foods consumed. Intake of pulses, nuts and seeds, and soy products increased in the 50/50 and Plant groups, while total, red, and processed meat intake decreased in the Plant group and increased in the Animal group (Fig. S1A). Meat intake remained stable in the 50/50 group, and dairy intake decreased with increasing plant-protein intake. Bray-Curtis distances between baseline and intervention food-group profiles were consequently greater in the 50/50 and Plant groups than in the Animal group (*p* < 0.001; Fig. S1B), showing that these diets differed more from participants’ habitual diets. Given the relevance of fiber intake to both CRC risk and the gut microbiota (Aune et al. 2011; O’Keefe et al. 2015), we separately examined the changes in dietary fiber compositions from 4-day food diaries. Total and insoluble fiber intake increased in the 50/50 and Plant groups, with the largest increases in the Plant group (*p* < 0.001; Fig. S1B). Soluble fiber intake did not change significantly in any group (*p* > 0.05).

Mantel tests were used to examine relationships among the dietary and omics datasets (Fig. 1B). Microbiota composition measured by 16S rRNA gene sequencing was strongly correlated with shotgun metagenomic taxonomic profiles (*r* = 0.76, *p* < 0.001) and was also significantly associated with metagenomic functional pathways, fecal BA profiles, nutrient intake, and food-group composition. Plasma BA profiles were weakly associated with nutrient intake (r = 0.08, p = 0.01) and marginally associated with metagenomic functional potential (r = 0.21, p = 0.06), but showed no significant association with microbiota composition measured by either sequencing approach (*p* > 0.05; Fig. 1B).

### Replacing animal-sourced with plant-sourced protein alters stool characteristics, restructures the gut microbiota, and lowers fecal NOCs

Stool characteristics differed across the three diets after the intervention (ANCOVA adjusted for baseline; Fig. 2A-C). Fecal pH decreased as the proportion of plant-sourced protein increased and was significantly lower after the Plant than the Animal diet (*p* < 0.01; Fig. 2A). Fecal weight was higher after the 50/50 diet than the Animal diet (*p* = 0.04) and tended to be higher after the Plant diet (*p* = 0.07; Fig. 2B). BSS was also higher after the Plant diet, indicating softer stools, with trends compared with both the 50/50 (*p* = 0.09) and Animal diets (*p* = 0.06; Fig. 2C). The combination of greater fecal output and softer stools is indicative of shorter colonic transit (Slavin et al. 1981).

**Figure 2.**
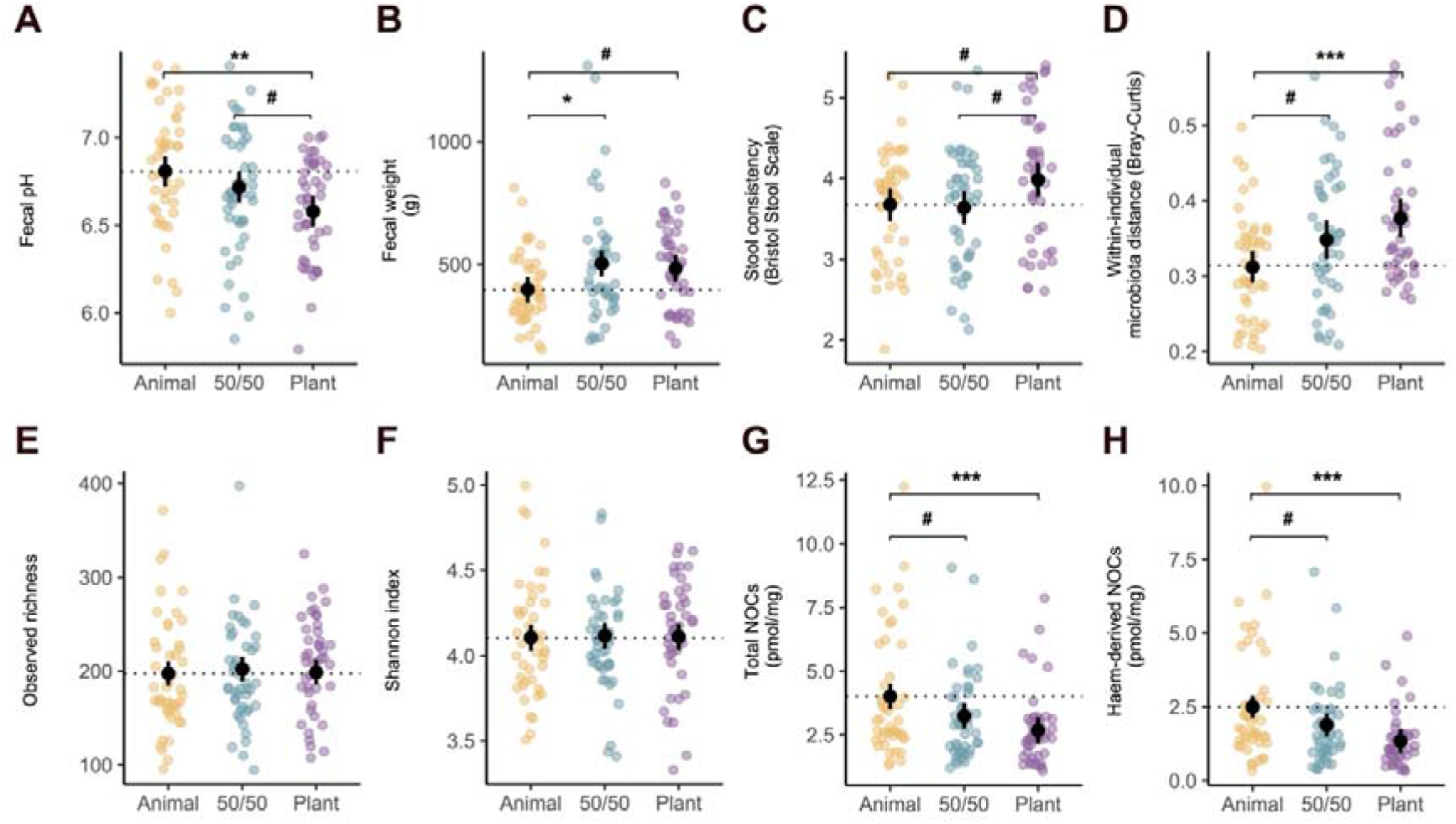
Effects of replacing animal-sourced with plant-sourced protein on stool characteristics, gut microbiota, and fecal *N*-nitroso compounds. (A-C) Fecal pH, fecal wet weight, and stool consistency assessed using the Bristol Stool Scale. **(D)** Within-individual Bray–Curtis dissimilarity between microbiota composition before and after the intervention. **(E,F)** Observed richness and Shannon diversity. **(G,H)** Fecal total and haem-derived N-nitroso compounds (NOCs). Differences between diets in panels A-C and E-H were assessed using ANCOVA adjusted for baseline values, followed by Bonferroni-adjusted pairwise comparisons. Between-group differences in within-individual Bray–Curtis dissimilarity **(D)** were assessed using Wilcoxon rank-sum tests. Black points and error bars indicate adjusted means and 95% confidence intervals in panels A–C and E–H, and group means with 95% confidence intervals in panel D. #*p* < 0.1, \**p* < 0.05, \*\**p* < 0.01, \*\*\**p* < 0.001.

Changes in gut microbiota composition also differed between diets. Within-participant Bray-Curtis dissimilarity between baseline and post-intervention samples was significantly greater after the Plant than the Animal diet (*p* < 0.001; Fig. 2D), while the 50/50 diet showed a weaker trend in the same direction (*p* = 0.07). Thus, participants assigned to the Plant diet underwent the largest overall change in microbiota composition. Within each diet group, PERMANOVA showed a significant pre-post shift after the Plant diet (*p* = 0.012), with no significant change after the 50/50 or Animal diets (*p* > 0.05; Fig. S1C). Microbiota α-diversity, measured by observed richness and Shannon diversity, did not differ among the three diets after the intervention (p > 0.05; Fig. 2E,F).

*N*-nitroso compounds (NOCs) can form endogenously in the gastrointestinal tract and include compounds with carcinogenic potential (Cross et al. 2010). Their site and mechanism of formation remain uncertain, although colonic nitrosation of protein-derived substrates, potentially involving nitrate-reducing bacteria, has been hypothesized (Loeppky 1994; Silvester et al. 1997; Lunn et al. 2007; Russell et al. 2011; Kobayashi 2018). Fecal total and heme-derived NOCs decreased with increasing replacement of animal-sourced with plant-sourced protein and were significantly lower after the Plant than the Animal diet (*p* < 0.01; Fig. 2G,H).

### A plant-protein-dominant diet reduces CRC-associated bile acids and potentially promotes bile acid detoxification

Bile acids (BAs) are synthesized from cholesterol in the liver, conjugated predominantly with glycine or taurine, and secreted into bile. Following food intake, they are released into the small intestine, where they facilitate lipid digestion and the absorption of fat-soluble vitamins. Most BAs are reabsorbed in the terminal ileum and returned to the liver through enterohepatic circulation, whereas approximately 5-10% reach the colon. There, gut microbes transform the residual BA pool through deconjugation, oxidation and epimerization, and 7α-dehydroxylation, generating secondary BAs (Cai et al. 2022). DCA and LCA are among the secondary BAs most consistently implicated in colorectal carcinogenesis; deconjugation of taurine-conjugated BAs releases taurine, which can elevate microbial H S production in the colon and has been linked to CRC (Kühn et al. 2020; Collins et al. 2023; Yang S et al. 2025). We therefore examined how increasing the proportion of plant-based relative to animal-sourced protein affected fecal and circulating BA profiles using targeted metabolomics, quantifying 20 BA species. In addition to individual BAs, we analyzed non-mutually exclusive BA classes defined by origin and metabolic modification, including total, primary, secondary, unconjugated, glycine-conjugated, taurine-conjugated, 12α-hydroxylated, sulfated, and glucuronidated BAs (see Method details).

Between-group analysis using linear mixed-effects models (LMEs) showed that fecal total, secondary, 12α-hydroxylated, and taurine-conjugated BAs were significantly lower after the Plant diet than after both the 50/50 and Animal diets (all *q* < 0.2; Fig. 3 and Fig. S2). The most pronounced difference was observed for taurine-conjugated BAs between the Plant and Animal groups (*q* < 0.001), with greater decreases in TCDCA, TDCA, and TCA contributing to this difference (Fig. S2). The decreases in the CRC-associated secondary BAs DCA and LCA were also greater after the Plant diet than after both the 50/50 and Animal diets (*q* < 0.2; Fig. S2). Plasma BA profiles showed fewer between-group differences (Fig. 3 and Fig. S2). Glucuronidated BAs increased more after the Plant diet than after the Animal diet (*q* = 0.007; Fig. 3), mainly reflecting greater increases in GCDCA-3G and GDCA-3G (Fig. S2). No significant differences in fecal or plasma BAs were detected between the 50/50 and Animal groups.

**Figure 3.**
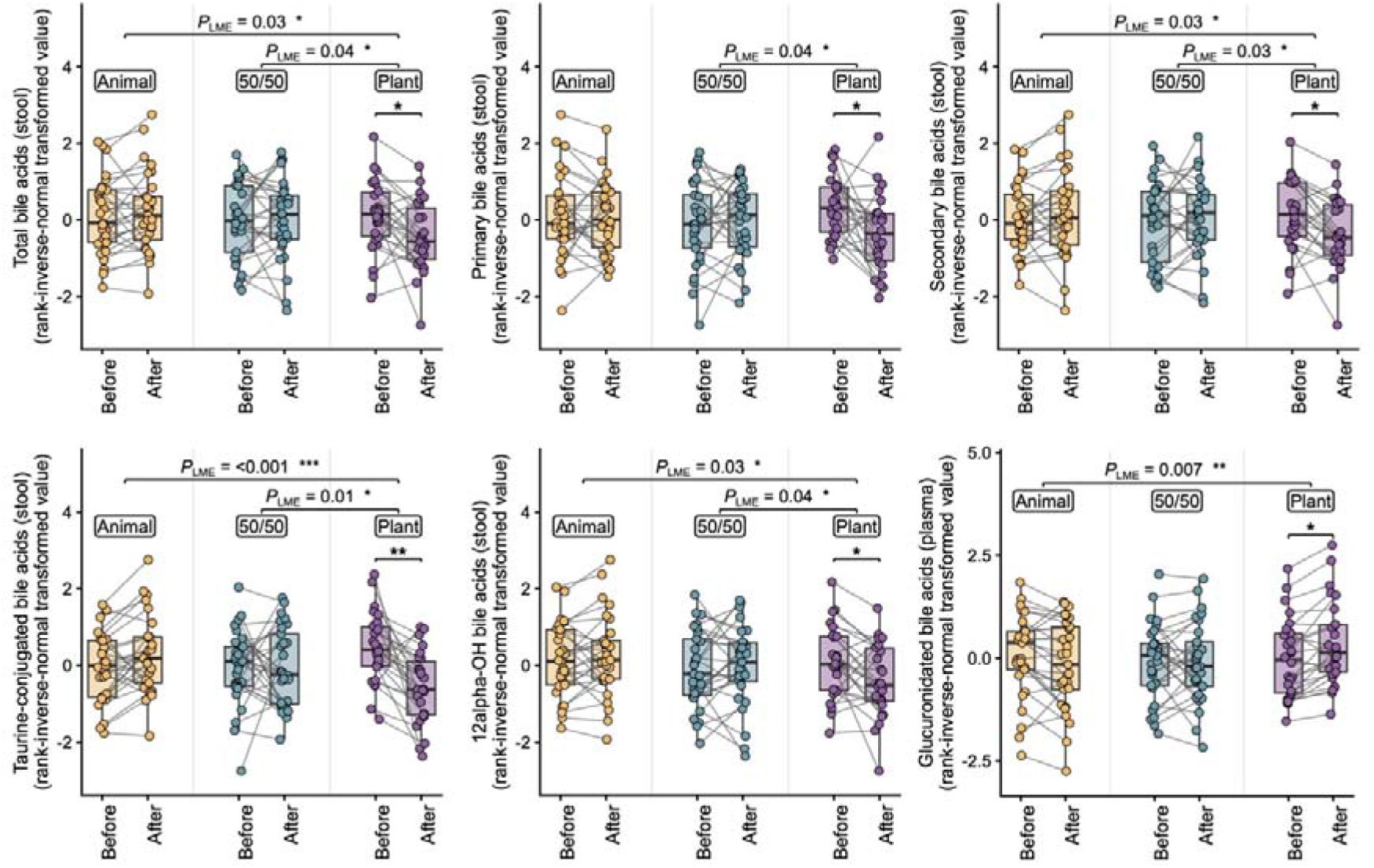
Effects of replacing animal-sourced with plant-sourced protein on fecal and plasma bile acids. Changes in fecal total, primary, secondary, taurine-conjugated, and 12α-hydroxylated bile acids and plasma glucuronidated bile acids across the intervention diets. Within-group pre–post changes were assessed using Wilcoxon signed-rank tests (*p* < 0.05, \**p* < 0.01, \*\**p* < 0.001), and differences in changes between diets were assessed using linear mixed-effects models (LME). Only bile acid groups with a significant difference in change over time in at least one pairwise diet comparison are shown. Brackets indicate significant pairwise diet × time interaction contrasts; displayed *P*_LME_ values are adjusted *p* values (*q* values). Gray lines connect paired samples from the same participant.

Within-group analyses showed that most major fecal BA classes decreased significantly from baseline in the Plant group, including total, primary, secondary, unconjugated, glycine- and taurine-conjugated, and 12α-hydroxylated BAs, whereas no significant decreases were observed in the 50/50 or Animal groups (Wilcoxon signed-rank test, all *q* < 0.2; Fig. S2). Plasma glucuronidated BAs also increased significantly from baseline in the Plant group, increasing the circulating pool of hydrophilic phase II BA conjugates involved in detoxification and elimination (Barbier et al. 2009).

### A plant-protein-dominant diet enriches fiber-associated taxa and reduces asaccharolytic bacteria

Having established that the Plant diet produced the largest overall shift in microbiota composition, we next identified the taxa contributing to this change. Between-group analyses identified several taxa that responded specifically to the Plant diet. *Haemophilus*, dominated by *Haemophilus parainfluenzae*, increased significantly more after the Plant diet than after both the 50/50 and Animal diets (Fig. 4). This genus has frequently been associated with fiber-rich dietary patterns in cross-sectional studies (Asnicar F. et al. 2021; Ma et al. 2021; Seel et al. 2023). *Veillonella* also increased after the Plant diet. Both *Haemophilus* and *Veillonella* are common inhabitants of the upper gastrointestinal tract and are sensitive to bile acids; their increase may therefore be related to the lower fecal BA levels after the Plant diet (Loomba et al. 2021; Lancaster et al. 2022). *Ruminococcaceae* UCG-013 also showed a greater increase after the Plant diet than after either of the other diets (Fig. 4), in line with previous associations with barley and legume intake (Matsuoka et al. 2022; Vauhkonen 2022). *Roseburia*, which also increased after the Plant diet, is a common responder to plant-rich diets and includes flagellated species capable of attaching to and degrading a range of dietary polysaccharides (Scott et al. 2011).

**Figure 4.**
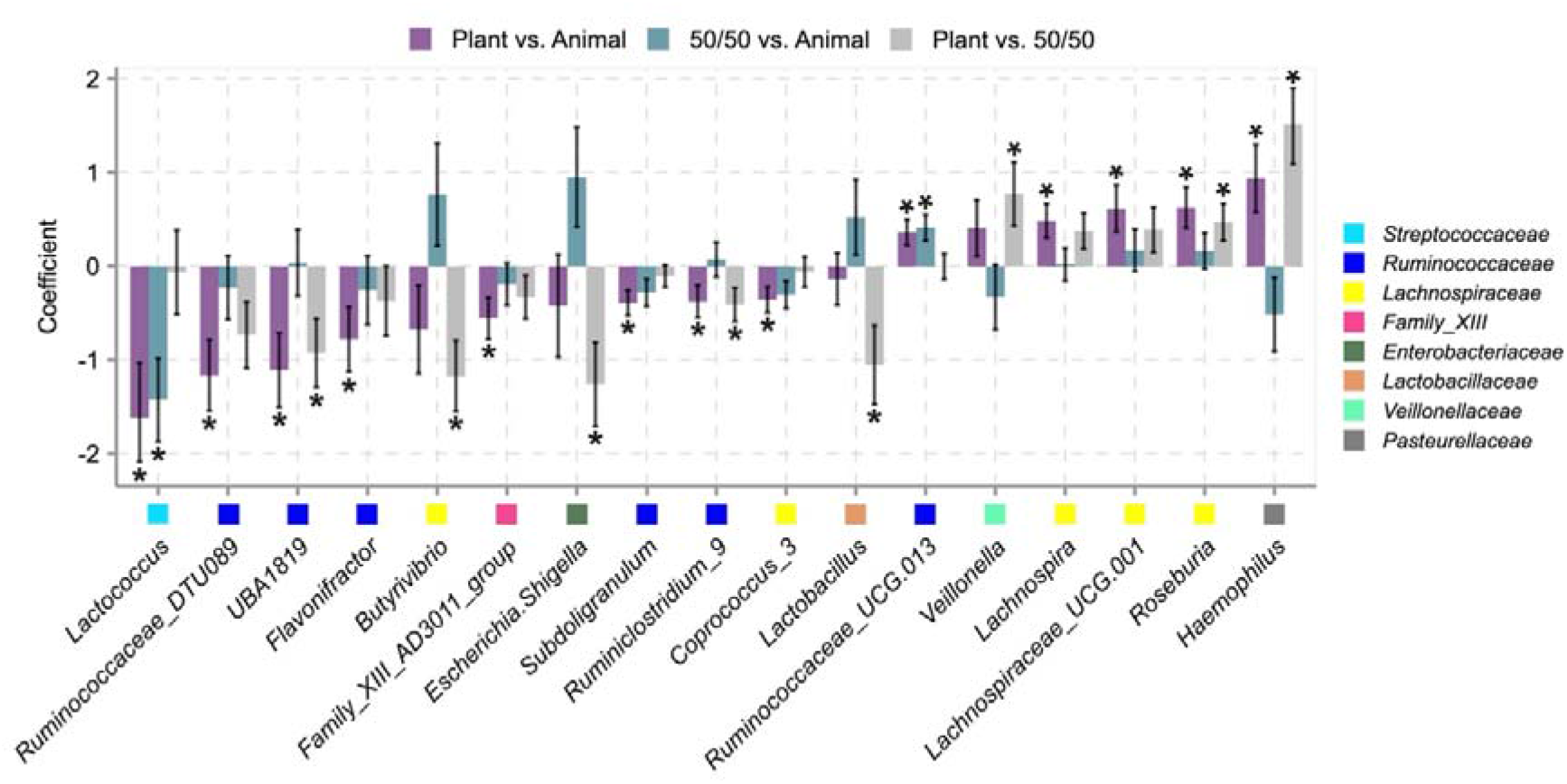
Effects of replacing animal-sourced with plant-sourced protein on gut bacterial genera. Bar plots show between-group differences in changes in genus-level relative abundance estimated using linear mixed-effects models (LME). Coefficients represent pairwise diet × time interaction contrasts. Error bars represent the standard errors of the estimated coefficients. Only genera with a significant difference (*q* < 0.2) in at least one comparison are shown. Positive coefficients indicate relative enrichment over time in the first diet of each contrast compared with the second, whereas negative coefficients indicate relative depletion. \**q* < 0.2.

Within-group analyses corroborated a much broader taxonomic response to the Plant diet. Of the 106 genera retained after filtering, 30% changed significantly in relative abundance (Wilcoxon signed-rank test, *q* < 0.2; Fig. S3). After the 50/50 diet, only *Lactococcus* decreased significantly, and no genus changed significantly after the Animal diet. Genera that increased after the Plant diet included *Lachnospiraceae* UCG-001, *Lachnospiraceae* UCG-004, *Lachnospira*, *Roseburia*, and *Butyricicoccus*, taxa repeatedly associated with fiber-rich Mediterranean and non-industrialized dietary patterns (Meslier et al. 2020; Zhu et al. 2020; Bourdeau-Julien et al. 2023; Li F et al. 2025). Among the taxa that decreased were several members of *Ruminococcaceae* and Family XIII AD3011, together with dairy-associated *Streptococcaceae* (Fackelmann et al. 2025). Many of the reduced *Ruminococcaceae* and Family XIII AD3011 taxa are asaccharolytic and have been associated with protein fermentation and production of ammonium and indolic metabolites *in vitro* (Amaretti et al. 2019). Shotgun metagenomics resolved these changes to species level, with *Bittarella massiliensis*, *Anaeromassilibacillus* sp. An250, and *Ruthenibacterium lactatiformans* among the species showing the largest decreases after the Plant diet (Table S1).

We also estimated bacterial growth rates from the shotgun metagenomes using peak-to-trough ratios, which compare sequencing coverage near the origin and terminus of replication (Emiola and Oh 2018). Inferred growth rates increased significantly in 10 species after the Plant diet (Wilcoxon signed-rank test, *q* < 0.2; Fig. S4); nine belonged to *Lachnospiraceae*, including *Roseburia inulinivorans*. After the Animal diet, inferred growth rates increased in three species, including *Alistipes putredinis*, a species associated with animal-based diets (David et al. 2014; Fackelmann et al. 2025) and reported to be enriched in CRC (Janney et al. 2020). Thus, the Plant diet increased both the relative abundance and growth of several fiber-associated bacteria while reducing taxa with predominantly asaccharolytic metabolism.

### A plant-protein-dominant diet increases plant-carbohydrate degradation potential and reduces methanogenic potential

We examined whether the taxonomic changes were accompanied by changes in microbial functional potential. Gut Metabolic Modules (GMMs) were used to summarize KEGG orthologues (KOs) into defined microbial metabolic pathways (Vieira-Silva et al. 2016). Compared with the Animal diet, the Plant diet increased GMMs for degradation of pectin, rhamnose, and melibiose, as well as sulfate reduction (LME, *q* < 0.2; Fig. 5A). Increased carbohydrate-degradation potential has also been reported in individuals consuming vegetarian, vegan, or Mediterranean diets (Wang DD et al. 2021; Fackelmann et al. 2025). In contrast, GMMs for degradation of the amino acids leucine and lysine were relatively depleted after the Plant diet. Putrescine degradation also decreased; putrescine is a microbial polyamine involved in colonic epithelial renewal. GMMs for lactose and galactose degradation were similarly reduced, in line with the lower dairy intake in the Plant group (LME, all *q* < 0.2; Fig. 5A). Notably, two GMMs related to methanogenesis showed among the strongest reductions after the Plant diet relative to the Animal diet (LME, *q* < 0.2; Fig. 5A).

**Figure 5.**
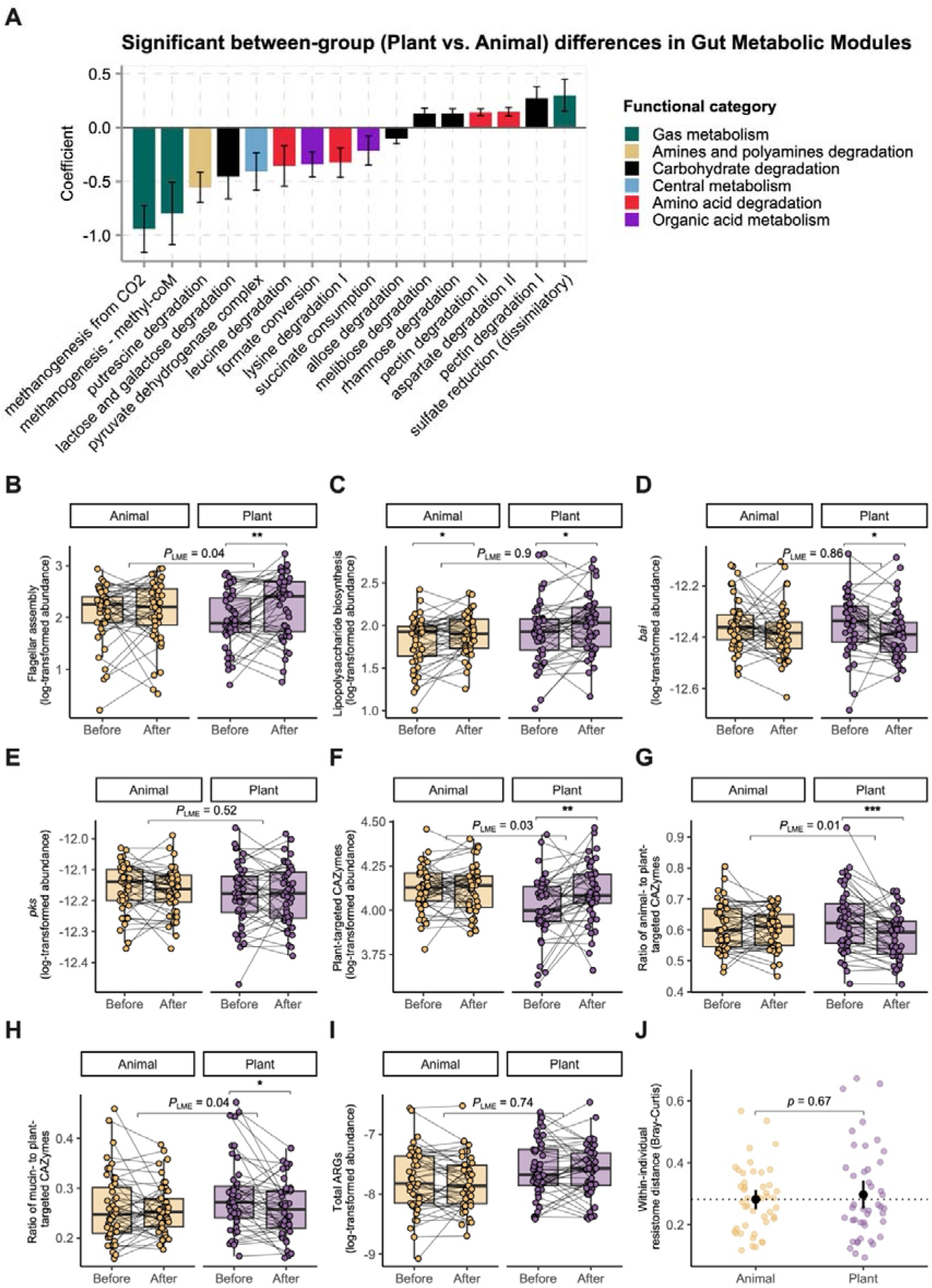
Effects of replacing animal-sourced with plant-sourced protein on gut microbial functional potential. **(A)** Between-group differences in pre–post changes in Gut Metabolic Modules (GMMs) between the Plant and Animal diets, estimated using linear mixed-effects models (LME). Error bars represent the standard errors of the estimated coefficients. Only GMMs with a significant diet × time interaction are shown (*q* < 0.2). Positive coefficients indicate relative enrichment over time in the Plant diet compared with the Animal diet, whereas negative coefficients indicate relative depletion. Bars are colored by functional category. **(B–I)** Changes in flagellar assembly genes **(B)**, lipopolysaccharide biosynthesis genes **(C)**, *bai* genes **(D)**, *pks* genes **(E)**, plant-targeted carbohydrate-active enzymes (CAZymes) **(F)**, the ratio of animal-to plant-targeted CAZymes **(G)**, the ratio of mucin- to plant-targeted CAZymes **(H)**, and total antimicrobial resistance genes (ARGs) **(I)**. Gray lines connect paired samples from the same participant. Within-group changes were assessed using Wilcoxon signed-rank tests (*p* < 0.05, \**p* < 0.01, \*\**p* < 0.001), and differences in pre–post changes between diets were assessed using LME models; displayed *P*_LME_ values are adjusted *p* values (*q* values). **(J)** Within-individual Bray-Curtis dissimilarity of ARG composition over the intervention in the Animal and Plant groups.

In addition to GMMs, we examined additional functional modules that could support microbial adaptation to the greater supply of plant-derived carbohydrates and the faster-transit intestinal environment suggested by the stool changes (Fig. 5B–H). These included flagellar motility, which can facilitate nutrient access and help bacteria resist luminal washout (Friedlander et al. 2013), and carbohydrate-active enzymes (CAZymes) involved in the degradation of complex dietary and host carbohydrates (Wardman et al. 2022). Flagellar assembly genes increased significantly after the Plant diet (Wilcoxon signed-rank test, *p* < 0.01), with a greater increase than after the Animal diet (LME, *q* = 0.04; Fig. 5B). CAZymes targeting plant carbohydrates also increased, accompanied by decreases in the ratios of animal- to plant-carbohydrate CAZymes and mucin- to plant-carbohydrate CAZymes (Fig. 5F–H). These changes support a shift in carbohydrate utilization toward plant-derived substrates and away from animal-derived carbohydrates and host mucin.

We also examined microbial genes and pathways previously linked to CRC, including lipopolysaccharide (LPS) biosynthesis (Li Q et al. 2023), *Fusobacterium nucleatum* adhesion protein A (*fadA*), *Bacteroides fragilis* enterotoxin (*bft*), the *pks* genomic island responsible for colibactin production in some *Escherichia coli* strains, and the *bai* operon involved in secondary bile acid conversion (Wirbel et al. 2019). LPS biosynthesis potential increased modestly after both the Plant and Animal diets (Wilcoxon signed-rank test, *p* < 0.05; Fig. 5C), but the changes did not differ significantly between diets (LME, *q* > 0.2). *bai* gene abundance decreased after the Plant diet (Wilcoxon signed-rank test, *p* < 0.05; Fig. 5D), whereas the changes in both *bai* and *pks* abundance did not differ significantly between the Plant and Animal diets (LME, *q* > 0.2; Fig. 5D,E). Neither *fadA* nor *bft* was detected in this healthy cohort.

Finally, we examined antimicrobial resistance genes (ARGs), for which associations with diet have been reported but remain inconsistent across cross-sectional studies (Oliver et al. 2022; Stege et al. 2022; Pärnänen et al. 2024). Neither total ARG load nor ARG composition changed significantly after the Plant or Animal diet (Fig. 5I,J).

### Insoluble fiber intake is strongly associated with gut microbiota shifts and lower fecal NOCs

The substantial changes in food intake during the intervention allowed us to examine which dietary components were most closely associated with microbiota restructuring. Across all participants, the magnitude of microbiota change was significantly associated with Bray-Curtis food distance from the baseline diet and with changes in total and insoluble fiber intake (all *p* < 0.01; Fig. 6A), but not with soluble fiber intake (*p* > 0.05). No significant associations were found for changes in total energy, protein, carbohydrate, or fat intake (*p* > 0.05; Fig. S5A).

**Figure 6.**
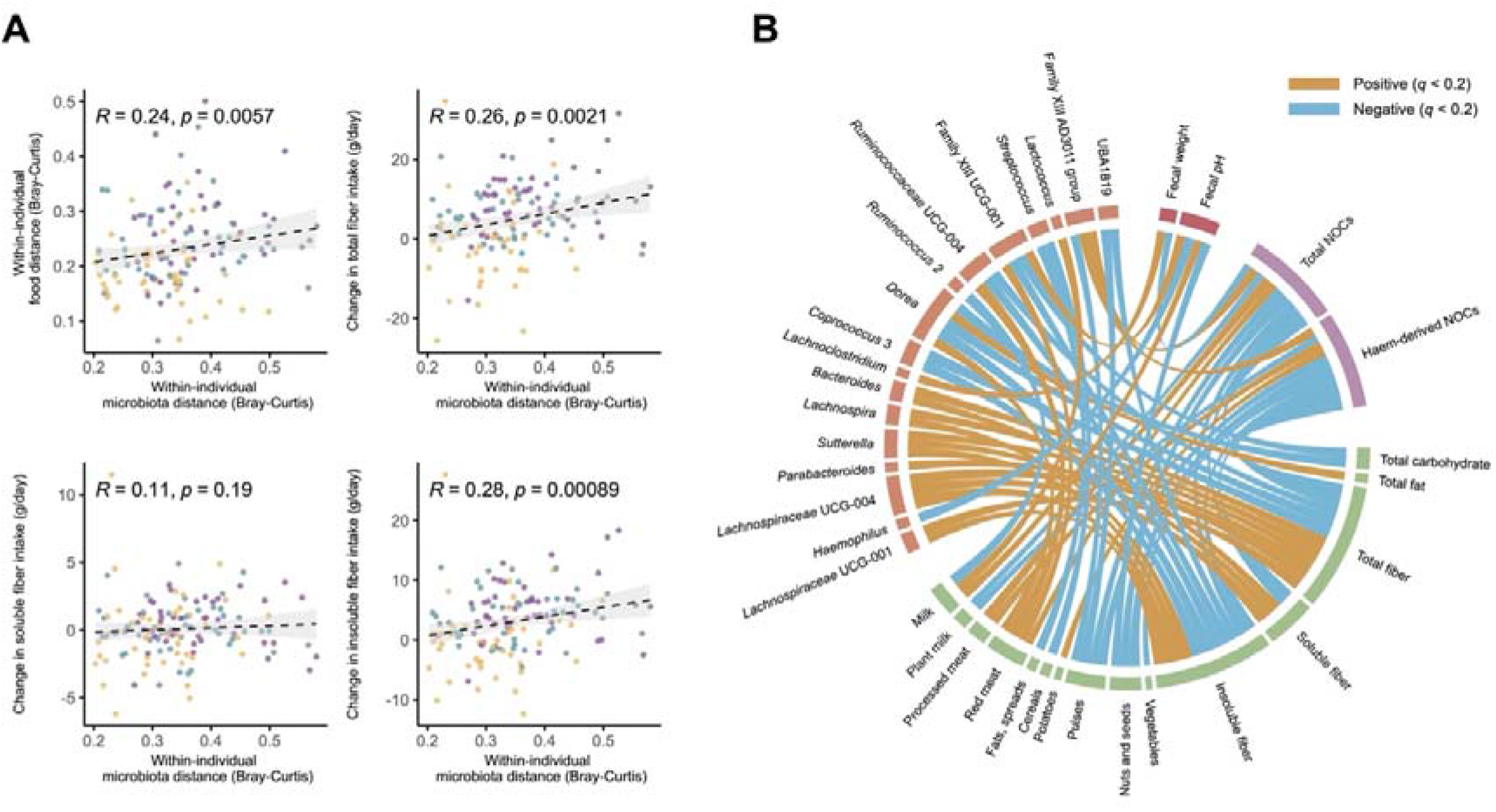
Associations between dietary change, gut microbiota restructuring, and fecal NOCs. **(A)** Correlations between within-individual gut microbiota change, quantified as Bray-Curtis dissimilarity between baseline and post-intervention samples, and the corresponding change in diet, quantified as Bray-Curtis food distance from baseline, as well as changes in total, soluble, and insoluble fiber intake. Each point represents one participant; colors denote intervention groups. Dashed lines show linear fits with 95% confidence bands. **(B)** Chord diagram summarizing significant Spearman correlations among genus-level relative abundance, dietary intake, fecal *N*-nitroso compounds (NOCs), and stool characteristics during the intervention. Only variables with at least one significant association are shown (*q* < 0.2). Orange and blue connectors indicate positive and negative correlations, respectively.

We next examined associations among genus-level relative abundance, dietary intake, fecal NOCs, and stool characteristics using Spearman correlations (Fig. 6B). Fecal NOCs and dietary fiber showed the largest numbers of significant associations, with 22 and 18 correlations, respectively. NOC levels were positively associated with red and processed meat intake and negatively associated with dietary fiber, particularly fiber from pulses and nuts/seeds. In contrast, associations with individual bacterial taxa were limited. Family XIII AD3011 (*Anaerovoracaceae*, Clostridia) was the only taxon positively associated with fecal NOCs and was itself strongly associated with methanogenesis (*r* = 0.39, *p* < 0.001; Fig. S5B). Changes in fecal NOCs were not associated with metagenomic nitrate-reduction potential (*r* = −0.14, *p* = 0.20), but were associated with methanogenesis (*r* = 0.32, *p* = 0.002; Fig. S5B).

The taxa associated with insoluble fiber also closely reflected the microbiota changes induced by the Plant diet. *Lachnospiraceae* UCG-001, *Lachnospiraceae* UCG-004, and *Lachnospira*, all enriched after the Plant diet, correlated positively with fiber intake, most strongly with insoluble fiber (Fig. 6B). By contrast, insoluble fiber was negatively associated with *Coprococcus* 3, predominantly represented by *Coprococcus comes*, as well as *Dorea*, *Ruminococcaceae* UCG-004, Family XIII UCG-001, and Family XIII AD3011; all of these taxa decreased in relative abundance after the Plant diet.

Overall, insoluble fiber showed the strongest and most consistent associations with microbiota restructuring and lower fecal NOCs, whereas fecal NOC variation was more closely related to dietary intake than to the abundance or functional potential of nitrate-reducing microbes.

### Methanogen carriage reflects an unfavorable colonic environment and predicts weaker responses to the Plant diet

Because methanogenesis decreased after the Plant diet and gut methanogens have been associated with CRC (Piqué et al. 1984; Hibberd et al. 2017; Coker et al. 2020; Mathlouthi et al. 2023; Li T et al. 2024), we tested whether pre-intervention methanogen carriage was related to intestinal characteristics and subsequent response to the intervention. *Methanobrevibacter*, the dominant methanogenic genus in the human gut, was detected by 16S rRNA gene sequencing in 44 of 136 participants (32%) at baseline, in agreement with previous qPCR-based estimates (Lahti et al. 2014). These participants were classified as methanogen carriers.

Carriers had a distinct baseline microbiota composition from non-carriers (PERMANOVA, *p* = 0.001; Fig. 7A). *Christensenellaceae* and *Ruminococcaceae*, which commonly form syntrophic associations with *Methanobrevibacter* (Kumpitsch et al. 2021; van der Vossen et al. 2023), were enriched in carriers, together with *Akkermansia*, *Butyricimonas*, *Desulfovibrio*, *Oxalobacter*, and Family XIII AD3011 (Fig. S6A), taxa previously associated with slower colonic transit (Müller et al. 2020; Procházková et al. 2023). *Faecalibacterium*, *Ruminococcaceae* UCG-013, *Roseburia*, *Lachnoclostridium*, and *Erysipelotrichaceae* UCG-003 were depleted (Fig. S6A). Metagenomic analysis showed greater amino acid-degradation potential and lower carbohydrate-metabolism potential in carriers (Fig. S6B), resembling the shift toward proteolytic metabolism reported with prolonged colonic transit (Procházková et al. 2023) and in CRC-associated microbiomes (Wirbel et al. 2019). Carriers also had lower fecal weight and BSS, indicating harder stools, together with higher microbiota α-diversity (all *p* < 0.05; Fig. 7B). Fecal pH did not differ between groups, whereas baseline fecal NOC concentrations were significantly higher in carriers (*p* < 0.01; Fig. 7B). No significant differences were detected in sex, age, BMI, or baseline intake of major food groups or nutrients between carriers and non-carriers (all p > 0.05).

**Figure 7.**
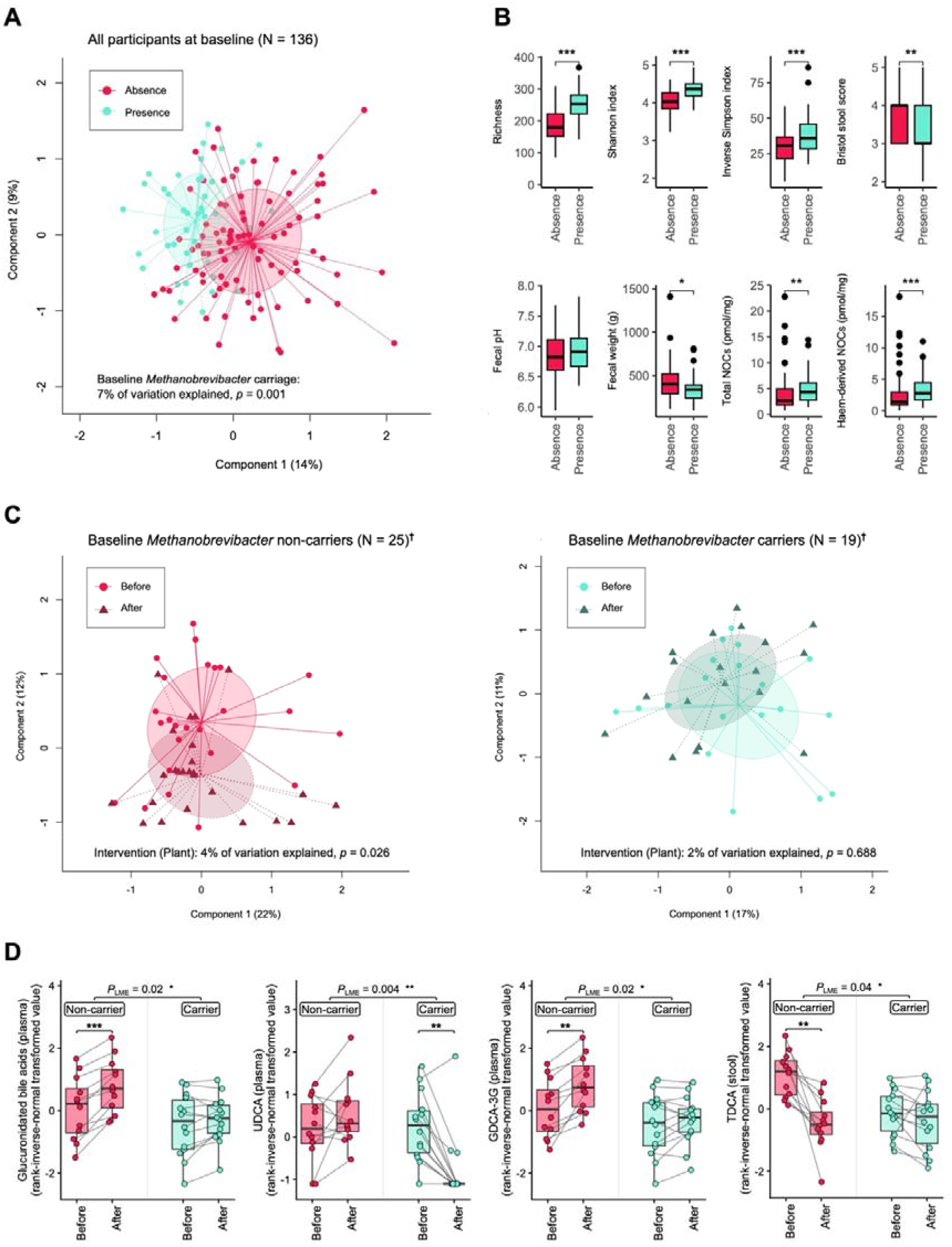
Baseline *Methanobrevibacter* carriage is associated with gut microbiota characteristics and differential responses to the Plant diet. **(A)** Principal coordinates analysis (PCoA) based on Bray-Curtis dissimilarity showing baseline microbiota composition in *Methanobrevibacter* non-carriers and carriers (N = 136). Carriage status explained 7% of the variation in microbiota composition (PERMANOVA, *p* = 0.001). **(B)** Baseline differences between non-carriers and carriers in microbiota α-diversity, Bristol Stool Scale score, fecal pH, fecal wet weight, and fecal total and haem-derived *N*-nitroso compounds (NOCs). Between-group differences were assessed using Wilcoxon rank-sum tests. **(C)** PCoA showing microbiota composition before and after the Plant diet separately in baseline non-carriers (n = 25) and carriers (n = 19). The Plant diet was associated with a significant change in microbiota composition in non-carriers (PERMANOVA, 4% of variation explained, *p* = 0.026), but not in carriers (2%, *p* = 0.688). **(D)** Changes in plasma total glucuronidated bile acids, plasma UDCA, plasma GDCA-3G, and fecal TDCA after the Plant diet according to baseline *Methanobrevibacter* carriage. Gray lines connect paired samples from the same participant. Within-group pre– post changes were assessed using Wilcoxon signed-rank tests, and differences in these changes between non-carriers and carriers were assessed using linear mixed-effects models testing the carriage status × time interaction; displayed *P*_LME_ values are adjusted *p* values (*q* values). \**p* < 0.05, \*\**p* < 0.01, \*\*\**p* < 0.001. †Number of participants with paired microbiota profiles before and after the Plant diet.

Methanogen carriage was highly stable during the intervention, with more than 90% of participants retaining their baseline status over 12 weeks irrespective of diet (Fig. S7A). *Methanobrevibacter* abundance nevertheless decreased significantly after the 50/50 and Plant diets (Wilcoxon signed-rank test, *p* < 0.05; Fig. S7B), with a greater reduction after the Plant than the Animal diet (LME, *p* = 0.03). Moreover, methanogenesis potential decreased more after the Plant than the Animal diet (LME, *p* = 0.04; Fig. S7B).

The Plant diet produced a significant shift in microbiota composition in non-carriers (PERMANOVA, *p* = 0.026), whereas no significant shift was detected in carriers (*p* = 0.688; Fig. 7C). Within-group taxonomic analyses identified changes in 24 genera in non-carriers compared with 11 in carriers (Fig. S7C). Bile acid responses also differed according to pre-intervention methanogen carriage. The decrease in fecal TDCA was greater in non-carriers than in carriers (LME, *q* = 0.04; Fig. 7D and Fig. S8). Within non-carriers, TDCA, TCA, and TCDCA all decreased significantly from baseline, whereas none changed significantly in carriers (Fig. S8). Plasma total glucuronidated BAs also increased more in non-carriers (LME, *q* = 0.02), largely attributable to GDCA-3G (*q* = 0.02; Fig. 7D). Both total glucuronidated BAs and GDCA-3G increased significantly from baseline in non-carriers but not in carriers. The plasma UDCA response likewise differed between groups (LME, *q* = 0.004); UDCA, a secondary BA with proposed chemopreventive properties (Cai et al. 2022; Režen et al. 2022), decreased significantly only in carriers (Fig. 7D and Fig. S8). The change in fecal NOCs after the Plant diet did not differ significantly between carriers and non-carriers. Overall, methanogen carriers appeared to derive fewer benefits from the Plant diet, particularly in terms of microbiota restructuring and bile acid metabolism.

## Discussion

A recent systematic review identified only four randomized interventions of partial animal-to-plant protein replacement in healthy adults to date (Courtney et al. 2026). ScenoProt involved the most extensive dietary provision, with study foods accounting for approximately 80% of daily energy intake, compared with foods providing 25% of protein intake in BeanMan (Itkonen et al. 2024) and only the assigned meat or meat-alternative products in RE-MAP (Bianchi et al. 2022) and SWAP-MEAT (Crimarco et al. 2020). Of these, only ScenoProt and BeanMan used whole-food protein sources. Studying healthy adults under this level of dietary control allowed us to examine microbial and metabolic responses to protein-source replacement in realistic but strong exposure contrast settings, providing insights into the broader implications of the ongoing protein transition in developed economies. In this study, replacing animal-sourced protein with plant-sourced protein substantially increased total and insoluble fiber intake and restructured the gut microbiota toward fiber-degrading and bile acid-sensitive taxa, while methanogens and asaccharolytic taxa declined. The functional profile changed accordingly, with increased flagellar assembly potential, greater representation of CAZymes targeting plant carbohydrates, lower relative capacity for animal-carbohydrate and mucin degradation, and reduced proteolytic potential. These changes occurred alongside greater fecal bulk and lower fecal pH, suggesting that the altered colonic environment contributed to the observed microbial restructuring. The 70 E% plant-sourced protein diet also significantly reduced several CRC-associated luminal compounds, including DCA, LCA, taurine-conjugated BAs, and carcinogenic NOCs, with insoluble fiber intake showing the strongest association with the reduction in NOCs. Plasma glucuronidated BAs increased, suggesting enhanced phase II BA detoxification and greater formation of hydrophilic conjugates that can be more readily eliminated (Barbier et al. 2009). The magnitude of these responses differed according to baseline *Methanobrevibacter* carriage, with carriers showing weaker microbiota shifts and less favorable BA responses after the 70 E% plant-sourced protein diet.

The considerable increase in insoluble fiber intake with the Plant diet may help explain the changes in stool characteristics and part of the microbiota response. Recent studies in mice (Glenny et al. 2024) and dogs (Bhosle et al. 2025) reported larger microbiota shifts with insoluble than soluble fiber, suggesting that fermentability alone does not determine microbial responses to dietary fiber. Compared with readily fermented soluble fibers, many insoluble fibers are fermented more slowly and retain physical properties that increase fecal bulk and water retention and shorten colonic transit (McRorie and McKeown 2017). Consistent with this, the Plant diet reduced slow-growing *Methanobrevibacter* and slow-transit-associated taxa such as *Ruthenibacterium lactatiformans* (Roager et al. 2016; Müller et al. 2020; Asnicar Francesco et al. 2021). Methane slows down intestinal motility, while experimentally slowing colonic transit can induce methane excretion in initially methane-negative individuals, making a bidirectional relationship (El Oufir et al. 1996; Procházková et al. 2023). Our findings support the link between methanogenesis and slower transit with greater bacterial protein catabolism and accumulation of fecal carcinogens. Elevated methanogenesis has been reported in CRC and high-risk populations (Piqué et al. 1984; Kunkel et al. 2011; Li T et al. 2024), and a recent multi-cohort study found progressive enrichment of gut methanogens from healthy controls to adenoma and CRC independent of transit time, raising the possibility that methanogens may also contribute to colorectal carcinogenesis through mechanisms beyond altered gut transit (Li T et al. 2024).

Several whole-diet interventions rich in unprocessed or minimally processed plant foods have documented enrichment of motile, fiber-associated taxa such as *Lachnospiraceae* and *Roseburia* (Meslier et al. 2020; Zhu et al. 2020; Bourdeau-Julien et al. 2023; Li F et al. 2025). Flagellar motility may help these gut bacteria persist under faster luminal flow, whereas taxa favored by prolonged transit often rely more heavily on host-derived substrates such as mucin (McLoughlin et al. 2016). This could account for the concurrent increase in flagellar assembly potential and reduction in mucin-utilizing capacity in the microbiota after the Plant diet. Although flagellins are primary ligands for Toll-like receptor 5 (TLR5) that could trigger immune responses, the CRC relevance of increased flagellar assembly potential is less straightforward. Notably, several *Lachnospiraceae* produce silent flagellins that do not activate TLR5 downstream signaling (Clasen et al. 2023), while experimental studies have reported anti-proliferative effects of fecal flagellin (Nishijima et al. 2016) and protection of epithelial barrier function by *Roseburia* flagellin (Seo et al. 2020). The flagellin derived from certain *Roseburia* strains has, however, also been reported in the pre-colectomy microbiome of patients with hereditary CRC (Yan et al. 2020). Therefore, the source and activity of the flagellins matter more than total flagellar gene abundance and warrant further investigation.

Fecal pH also decreased after the Plant diet despite unchanged predicted SCFA-production capacity. Reduced proteolytic metabolism may have contributed by lowering production of alkaline metabolites such as ammonia (Amaretti et al. 2019). In fact, non-fermentable, insoluble cellulose has been shown to lower fecal pH and accelerate colonic transit more effectively than fermentable fiber such as pectin (Hillman et al. 1983). Since most harmful bacterial enzymes and methanogens operate optimally at neutral to mildly alkaline pH (McGarr et al. 2005), the lower colonic pH may provide an additional protective mechanism against CRC.

Fecal BA reductions after the Plant diet were similar to those reported in Mediterranean diet interventions (Ghosh et al. 2020; Meslier et al. 2020), in which animal protein intake is 50– 60% lower than in Western diets (Tosti et al. 2018). Unconjugated hydrophobic BAs such as DCA and LCA can act as detergents, disrupting cellular membranes and inducing cell-damage responses (Collins et al. 2023). DCA can also accelerate proteasomal degradation of p53 (Qiao et al. 2001), and has been shown to mediate Western diet-associated colonic tumorigenesis in animal models (Osswald et al. 2026). The marked reduction in taurine-conjugated BAs may partly reflect the lower intake of taurine-rich animal foods and would reduce the taurine released in the colon for microbial production of H S, a potent carcinogen (Ridlon et al. 2016). The reduction in 12α-hydroxylated BAs may also affect host metabolism, as these BAs promote intestinal lipid absorption in mouse models (Li R et al. 2021), while higher circulating concentrations are associated with insulin resistance in humans (Haeusler et al. 2013). Higher fecal output and faster colonic transit provide a plausible explanation for part of the observed BA response by increasing BA excretion and shortening the time available for microbial 7α-dehydroxylation, with the concurrent decrease in *bai* gene abundance after the Plant diet supporting reduced conversion to DCA and LCA (Wolf et al. 2022). Glucuronidation increases BA hydrophilicity and facilitates biliary and urinary elimination, whereas gut microbial β-glucuronidases can hydrolyze these conjugates and promote reabsorption of the released BAs (Pellock and Redinbo 2017; Boyle et al. 2026). Faster colonic transit may limit the opportunity for microbial deglucuronidation and reabsorption, favoring elimination of glucuronidated BAs. The relative contributions of host glucuronidation and microbial deglucuronidation to the increase in plasma BA glucuronides require further investigation.

NOCs can induce DNA alkylation, and a recent analysis of 900 CRC cases identified an alkylating mutational signature associated with red meat intake (Gurjao et al. 2021), linking the lower fecal NOCs in our study to a mechanism relevant to colorectal carcinogenesis. Fecal NOCs may originate directly from the diet or be formed endogenously, with microbial nitrate reduction thought to contribute to endogenous formation despite limited experimental evidence (Kobayashi 2018; Barrett et al. 2020; Loke et al. 2020; Armet et al. 2022). However, fecal NOC concentrations were not associated with nitrate-reduction potential or with taxa previously shown to form NOCs *in vitro* (Kobayashi 2018), providing little support for these hypothesized microbial determinants. Instead, NOCs were positively associated with red and processed meat intake and negatively associated with dietary fiber, particularly insoluble fiber. Together with the greater fecal output and faster transit after the Plant diet, this points to reduced colonic accumulation as one contributor to lower fecal NOCs. The positive association between NOCs and methanogenesis fits this interpretation, as methanogen carriage was associated with a slower-transit phenotype. It also argues against nitrate reduction as a major determinant of NOC formation in this cohort, given the reported competition between nitrate reduction and methanogenesis (Yang C et al. 2016). Of note, the Plant diet unexpectedly increased sulfate-reduction potential despite lower intake of sulfur-containing amino acids. Because methanogens and sulfate-reducing bacteria compete for H in the colon (Smith et al. 2019), the reduction in methanogenesis may have increased H availability for sulfate reduction. This response differs from the increased sulfur metabolism associated with sulfur-rich animal-based diets and CRC risk (Wang Y et al. 2021).

Methanogen carriers had lower carbohydrate-metabolism potential and higher α-diversity at baseline, two features that may contribute to their smaller microbiota shift after the Plant diet (Salonen et al. 2014). Methanogen-associated communities also differ in substrate use; for instance, methane producers and non-producers have been reported to rely on different taxa for cellulose degradation (Chassard et al. 2010). Remarkably, BA responses to the Plant diet appeared to depend on methanogen carriage. Fecal TDCA decreased more in non-carriers; higher circulating TDCA has been associated with increased CRC risk in prospective studies (Loftfield et al. 2022). Plasma UDCA decreased only in carriers, while UDCA treatment reduced recurrence of colorectal adenomas with high-grade dysplasia in a phase III trial (Alberts et al. 2005). Together, these findings support baseline methanogen carriage as a stratification factor for testing heterogeneity in response to plant-based diets in future CRC-prevention trials.

The study has several limitations. Fiber intake was classified as total, soluble, and insoluble fiber, which does not capture differences in carbohydrate structure and physicochemical properties that could affect microbial utilization (Gidley and Yakubov 2019). This may partly explain why soluble fiber intake remained unchanged despite increased microbial potential for pectin, rhamnose, and melibiose utilization after the Plant diet. More detailed characterization of fiber and oligosaccharide composition would help identify the dietary substrates underlying these functional changes. Participants had relatively high baseline fiber intake (28.7 g/day) compared with population averages in North America (17 g/day) and Europe (18-24 g/day) (McKeown et al. 2022), which should be considered when comparing microbiota responses across populations with different habitual fiber intakes (Holmes et al. 2022). Finally, our cohort consisted of healthy Finnish adults and was predominantly female; studies in more diverse populations will be needed to assess generalizability.

## Conclusions

Replacing animal-sourced with plant-sourced protein reduced fecal NOCs and BAs, lowered microbial proteolysis and fecal pH, and increased fecal output, changes that may lower CRC risk through multiple mechanisms. The marked increase in insoluble fiber likely contributed through effects on microbial substrate use and colonic transit, reducing the opportunity for potentially harmful compounds to accumulate or undergo microbial transformation. This is relevant as the protein transition increasingly relies on processed plant-protein foods, in which protein fractionation can reduce fiber content and alter its physicochemical properties (Toh et al. 2022; Wang and Jian 2023). Food development should therefore consider not only how much fiber is retained and how readily it is fermented, but also whether its physicochemical properties support fecal bulk and colonic transit, if processed plant-protein foods are to reproduce the favorable colonic effects observed with whole-food replacement.

## Data Availability

All data produced in the present work are contained in the manuscript

## Acknowledgements

We thank the trial volunteers for their dedication and commitment to the study protocol.

## Data availability

Raw shotgun metagenomic sequencing data have been deposited in the European Nucleotide Archive under accession number PRJEB111297, and raw amplicon sequencing data under accession number PRJEB123728. Both datasets are publicly available as of the date of publication. Additional information required to analyze the data reported in this paper is available from the lead contact upon reasonable request.

