## SUPPLEMENTARY FIGURES for "Replacing animal-sourced with plant-sourced proteins alters gut microbiota and biomarkers associated with colorectal cancer risk in healthy adults: A randomized feeding study"

**Figure S1. Dietary and gut microbiota changes following graded replacement of animal-sourced with plant-sourced protein.**

**Figure S2. Between- and within-group changes in fecal and plasma bile acid profiles following the dietary intervention.**

**Figure S3. Within-group taxonomic and functional changes in the gut microbiota after the intervention.**

**Figure S4. Changes in GRiD-estimated bacterial growth rates after the Animal and Plant diets.**

**Figure S5. Associations of macronutrient intake and microbial functional potential with microbiota change and fecal NOCs.**

**Figure S6. Baseline taxonomic and functional differences associated with *Methanobrevibacter* carriage**

**Figure S7. Stability of *Methanobrevibacter* carriage status and microbiota responses during the intervention.**

**Figure S8. Bile acid responses to the Plant diet according to baseline *Methanobrevibacter* carriage.**

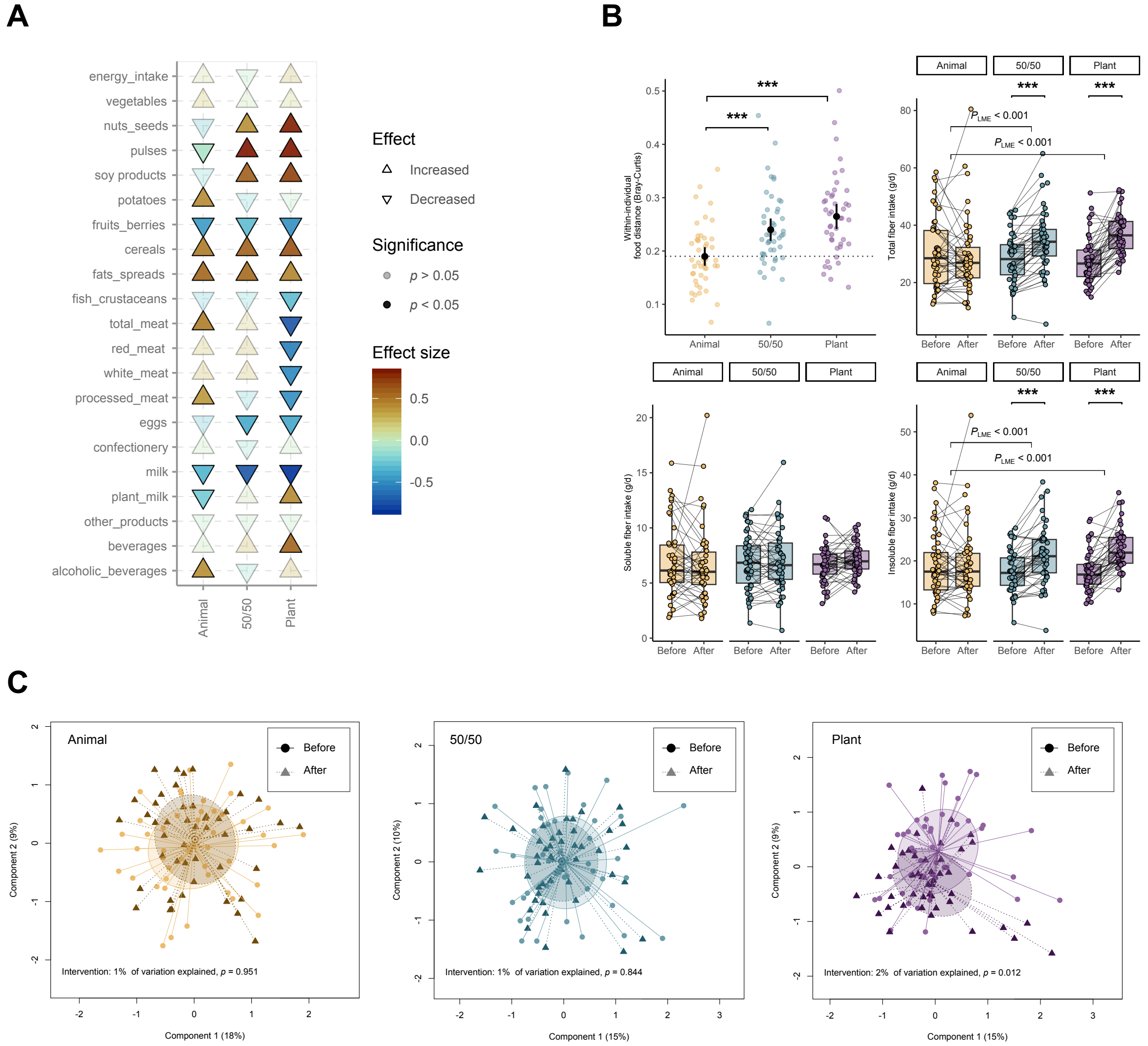


**Figure S1. Dietary and gut microbiota changes following graded replacement of animal-sourced with plant-sourced protein. (A)** Changes in food-group intake from baseline to the end of the intervention in the Animal, 50/50, and Plant groups. Upward and downward triangles indicate increased and decreased intake, respectively, and color indicates effect size. Solid triangles indicate significant within-group changes ($p < 0.05$). **(B)** Magnitude of dietary change from baseline, quantified as within-individual Bray-Curtis distance between food-group intake profiles, and changes in total, soluble, and insoluble fiber intake. Between-group differences in food distance were assessed using Wilcoxon rank-sum tests. Changes in fiber intake were assessed within groups using Wilcoxon signed-rank tests and between groups using linear mixed-effects models (LME). Asterisks indicate significant within-group changes (*$p < 0.05$, **$p < 0.01$, ***$p < 0.001$); brackets indicate between-group comparisons; displayed $P_{\text{LME}}$ values are adjusted $p$ values ($q$ values). Gray lines connect paired samples from the same participant. **(C)** Principal coordinates analysis (PCoA) based on Bray-Curtis dissimilarity showing gut microbiota composition before and after each intervention diet. The proportion of microbiota variation explained by the intervention and corresponding PERMANOVA $p$ value are shown for each group.

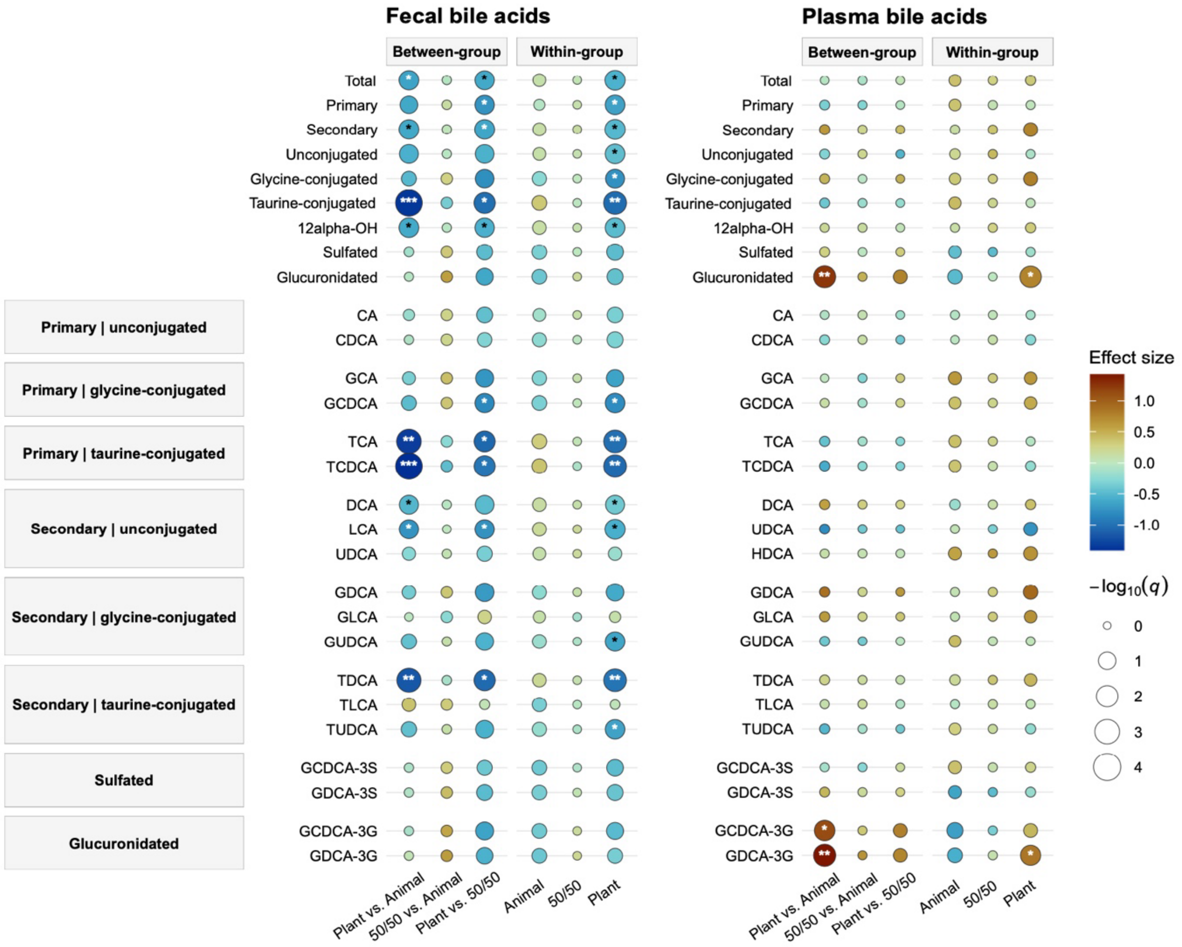


**Figure S2. Between- and within-group changes in fecal and plasma bile acid profiles following the dietary intervention.** Grouped bile acids are shown above the individual bile acids, which are arranged by primary/secondary origin and conjugation class. Between-group effects represent pairwise differences in pre-post change between diets estimated using linear mixed-effects models (diet × time interaction), whereas within-group effects represent pre-post changes within each diet. For between-group comparisons, positive effect sizes indicate relative enrichment over time in the first diet of each contrast compared with the second, whereas negative effect sizes indicate relative depletion. For within-group comparisons, positive and negative effect sizes indicate increases and decreases from baseline, respectively. Bubble color indicates effect size and bubble size represents −log10($q$), where $q$ denotes FDR-adjusted $p$ values. *$q < 0.05$, **$q < 0.01$, ***$q < 0.001$.

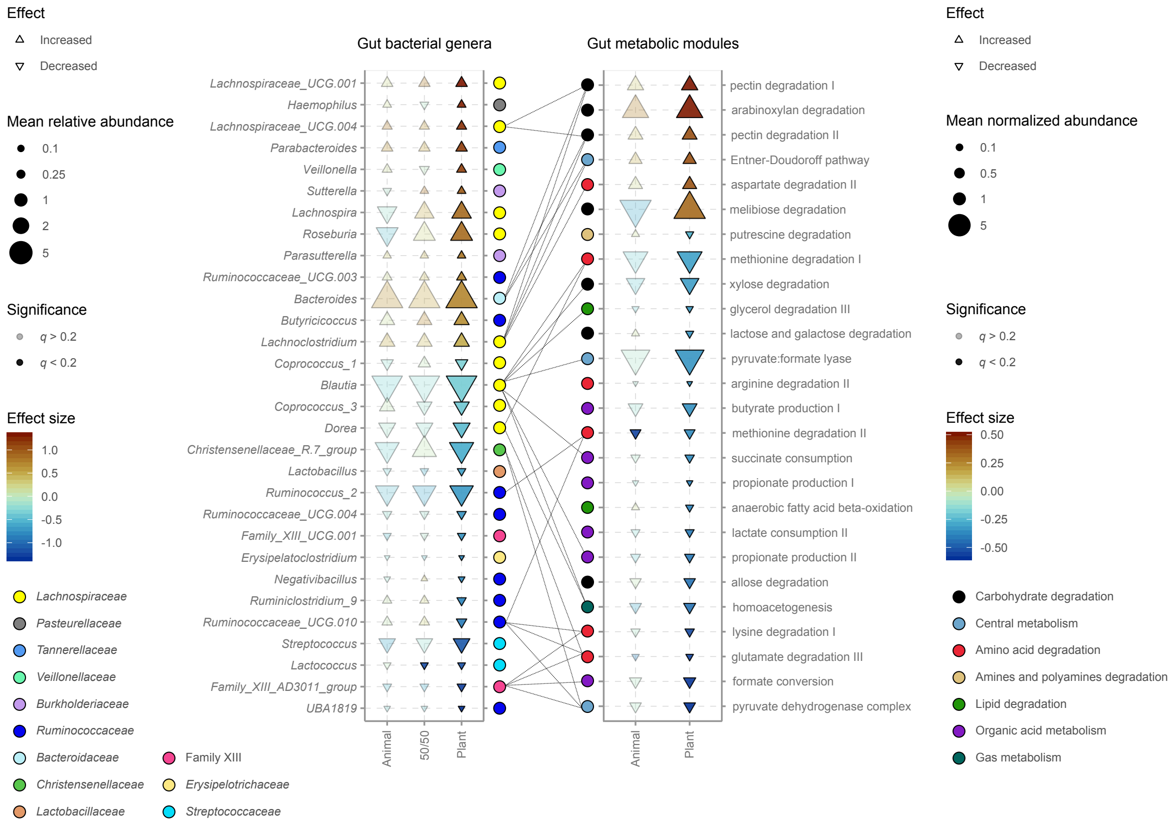


**Figure S3. Within-group taxonomic and functional changes in the gut microbiota after the intervention.** Changes in bacterial genera (left) and Gut Metabolic Modules (GMMs; right) from baseline to week 12 in the Animal, 50/50, and Plant groups. Upward and downward triangles indicate increases and decreases from baseline, respectively; triangle size represents mean abundance, color represents effect size, and solid triangles indicate significant changes ($q < 0.2$). Only genera or GMMs with a significant change in at least one intervention group are shown (Table S1). For bacterial genera, abundance is expressed as mean relative abundance; for GMMs, as mean normalized abundance. Colored circles indicate bacterial family and GMM functional category, respectively. Lines connect GMMs with potentially contributing genera based on Spearman correlations ($r > 0.4$; full associations in Table S2). Several carbohydrate-degradation GMMs correlated with *Bacteroides* and *Lachnoclostridium*, consistent with their known capacity for carbohydrate utilization and linking the taxonomic changes to the functional shifts observed during the intervention (PMID: 35166563; PMID: 36005398).

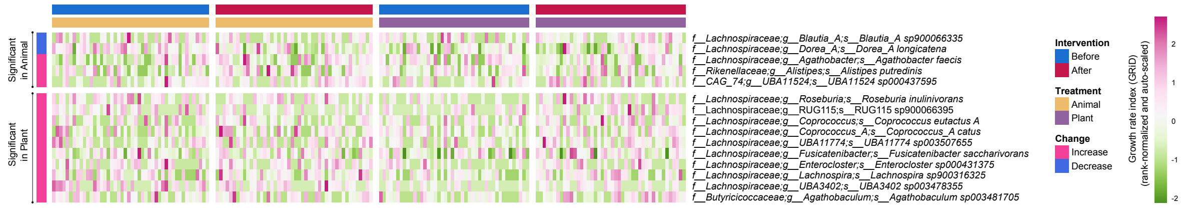


**Figure S4. Changes in GRiD-estimated bacterial growth rates after the Animal and Plant diets.** Heatmap showing bacterial species with a significant within-group change in inferred growth rate after either intervention diet (Wilcoxon signed-rank test, $q < 0.2$). Growth rates were estimated from shotgun metagenomes using the Growth Rate InDex (GRiD) algorithm, which infers bacterial replication from genome-wide differences in sequencing coverage associated with chromosome replication; higher GRiD values indicate faster inferred growth. Because bacterial growth rate does not necessarily track relative abundance, GRiD provides complementary information on taxa actively responding to the dietary intervention. Species are grouped according to the diet in which a significant change was detected. Columns represent individual samples before and after the intervention; heatmap colors indicate rank-normalized and auto-scaled GRiD values.

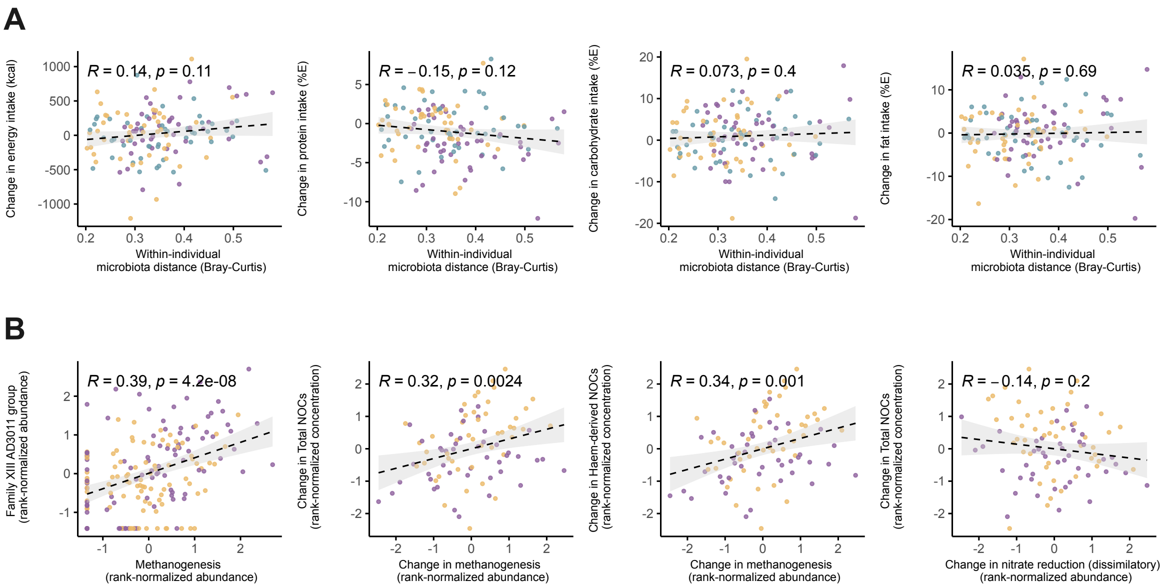


**Figure S5. Associations of macronutrient intake and microbial functional potential with microbiota change and fecal NOCs. (A)** Associations between within-individual Bray-Curtis microbiota dissimilarity and changes in energy, protein, carbohydrate, and fat intake over the intervention. None of these macronutrient changes was significantly associated with the extent of microbiota change. **(B)** Relationships linking methanogenesis to Family XIII AD3011 and fecal N-nitroso compounds (NOCs). Family XIII AD3011 abundance was positively associated with methanogenesis potential, and increases in methanogenesis were associated with increases in both total and haem-derived NOCs. By contrast, changes in nitrate-reduction potential were not associated with total NOCs. Spearman's *r* and corresponding *p* values are shown. Points represent individual participants and are colored by intervention group; dashed lines and shaded areas indicate fitted trends and 95% confidence intervals.

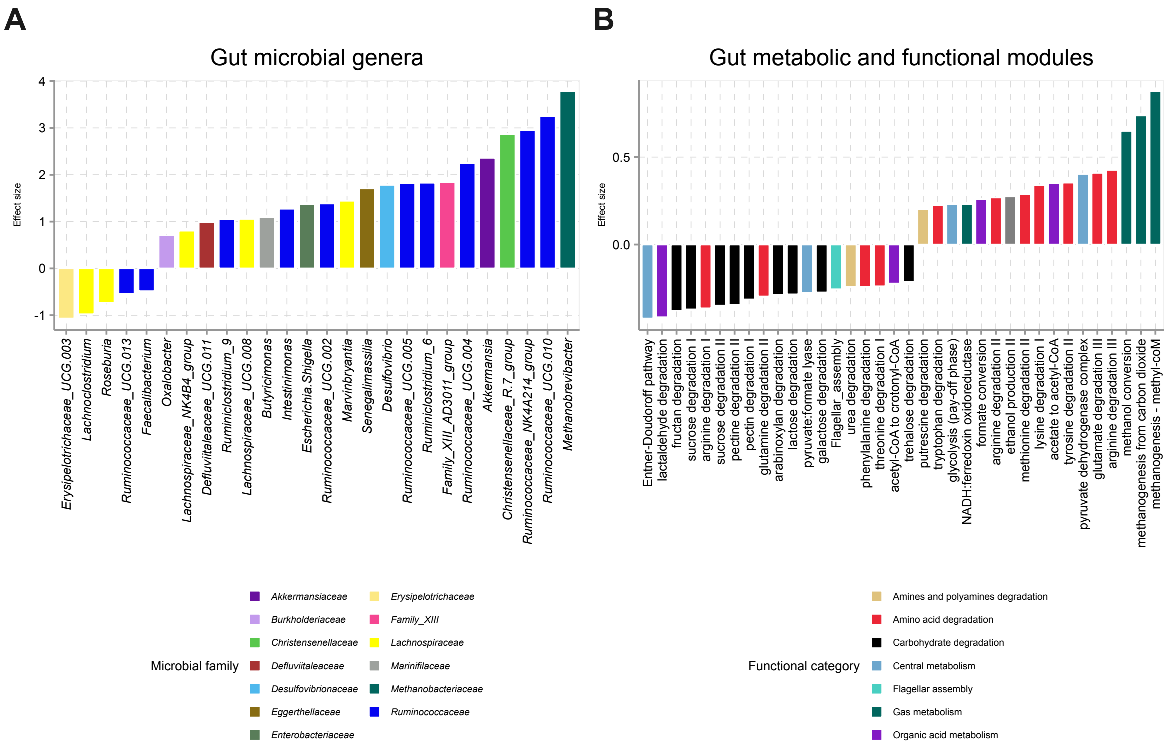


**Figure S6. Baseline taxonomic and functional differences associated with *Methanobrevibacter* carriage. (A)** Gut bacterial genera and **(B)** Gut Metabolic Modules (GMMs) differing between *Methanobrevibacter* carriers and non-carriers at baseline. Positive effect sizes indicate enrichment in carriers and negative effect sizes indicate depletion. Bars are colored by microbial family in (A) and functional category in (B). Only features with a significant between-group difference are shown ($q < 0.2$). Carriers were characterized by enrichment of *Methanobrevibacter* and several *Ruminococcaceae* taxa, together with higher methanogenesis and amino acid-degradation potential, whereas multiple carbohydrate-degradation pathways were depleted.

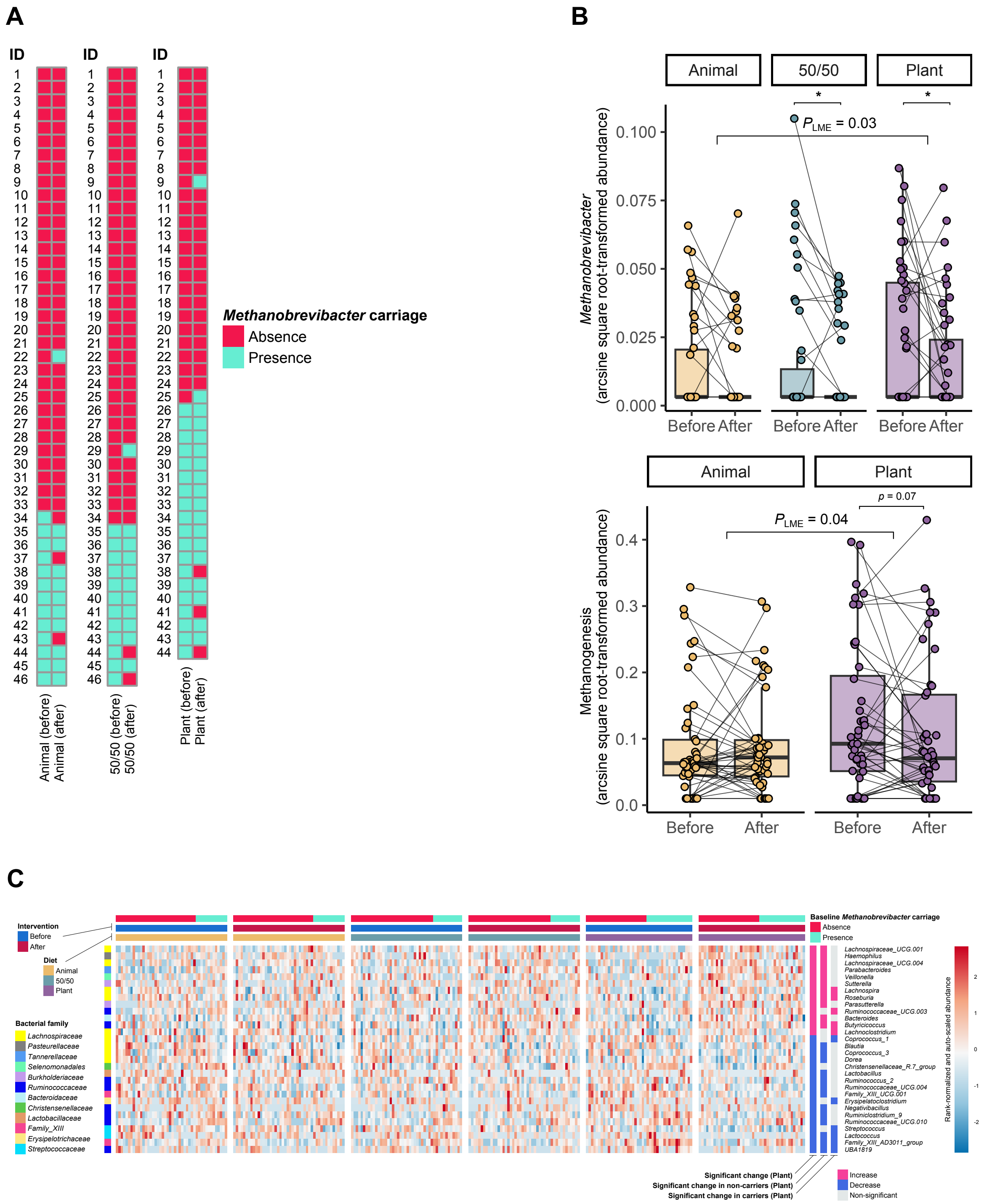


**Figure S7. Stability of *Methanobrevibacter* carriage status and microbiota responses during the intervention.** **(A)** *Methanobrevibacter* carriage before and after each intervention diet. Each row represents one participant; carriage status remained unchanged in >90% of participants over 12 weeks. **(B)** Changes in *Methanobrevibacter* relative abundance across the three diets (top) and metagenomic methanogenesis potential in the Animal and Plant groups (bottom). Gray lines connect paired samples. Within-group changes were assessed using Wilcoxon signed-rank tests ($*p < 0.05$, $**p < 0.01$, $***p < 0.001$), and differences in pre-post changes between diets using linear mixed-effects models (LME). *Methanobrevibacter* abundance decreased after the 50/50 and Plant diets, with a greater decrease after the Plant than the Animal diet; methanogenesis potential showed a similar between-diet difference. **(C)** Genus-level microbiota profiles before and after the intervention, annotated by diet and baseline *Methanobrevibacter* carriage. Rows show genera that changed significantly after the Plant diet overall or after stratification by baseline carriage status; side annotations indicate the direction and significance of these changes. More genera responded to the Plant diet in non-carriers than in carriers (24 vs. 11), consistent with the broader microbiota response observed in non-carriers. Heatmap colors represent rank-normalized and scaled relative abundance.

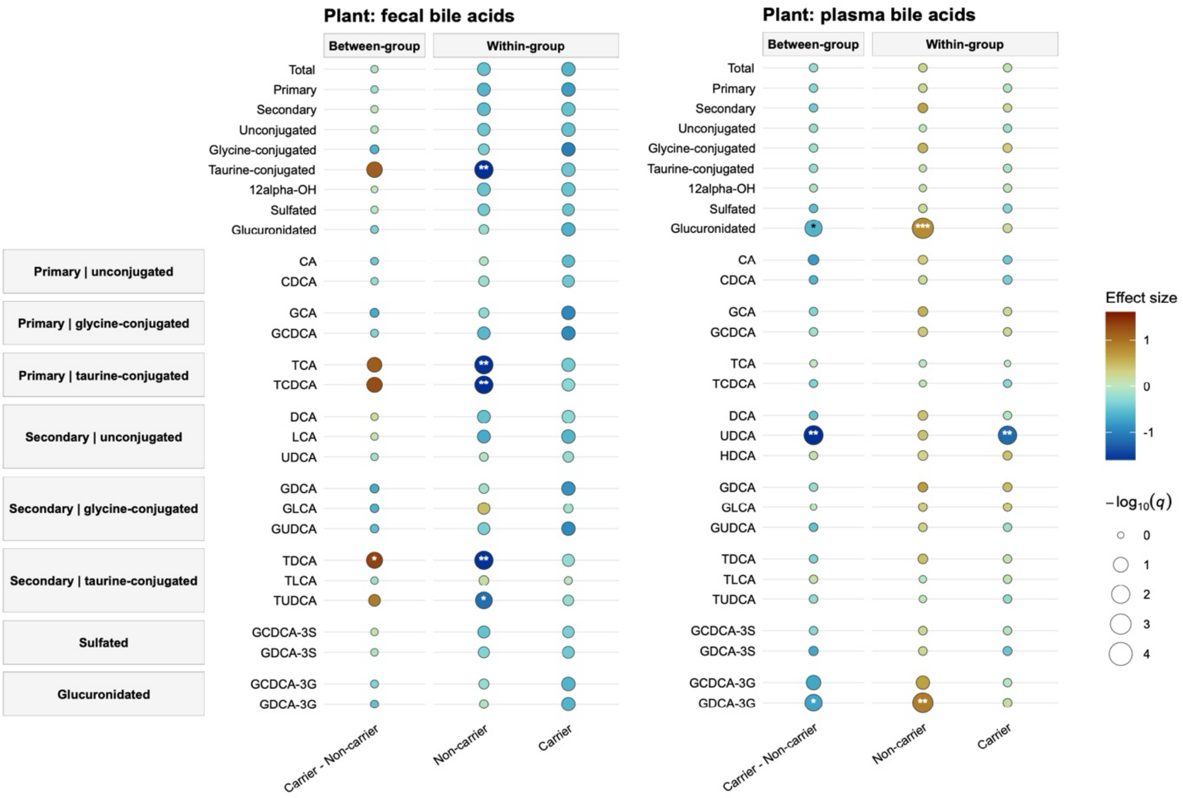


**Figure S8. Bile acid responses to the Plant diet according to baseline *Methanobrevibacter* carriage.** Fecal (left) and plasma (right) bile acid changes are shown for *Methanobrevibacter* non-carriers and carriers. Grouped bile acids are shown above individual bile acids, which are arranged according to primary or secondary origin and conjugation class. Between-group effects represent differences in pre-post changes between carriers and non-carriers estimated using linear mixed-effects models (carriage status × time interaction), whereas within-group effects represent pre-post changes separately in non-carriers and carriers. For the between-group comparison (carrier vs. non-carrier), positive effect sizes indicate relative enrichment over time in carriers and negative effect sizes indicate relative depletion. For within-group comparisons, positive and negative effect sizes indicate increases and decreases from baseline, respectively. Bubble color indicates effect size and bubble size represents −log10($q$), where $q$ denotes FDR-adjusted $p$ values. *$q < 0.05$, **$q < 0.01$, ***$q < 0.001$.
